# Prevalence and determinants of tobacco use among school-going adolescents in 53 African countries: evidence from Global Youth Tobacco Surveys

**DOI:** 10.1101/2024.05.24.24307910

**Authors:** Retselisitsoe Pokothoane, Terefe Gelibo Agerfa, Christus Cito Miderho, Noreen Dadirai Mdege

**Affiliations:** Development Gateway: an IREX Venture, Washington DC, United States; Research Unit on the Economics of Excisable Products (REEP), School of Economics, University of Cape Town, Cape Town, South Africa; Public Health, ICAP at Columbia University Mailman School of Public Health, Addis Ababa, Ethiopia; Département d’Agro-Vétérinaire, Institut Supérieur Pédagogique de Bukavu (ISP/Bukavu), BP 854 Bukavu, Democratic Republic of the Congo; Department of Health Sciences, University of York, York, United Kingdom; Centre for Research in Health and Development, York, United Kingdom

## Abstract

**Introduction:** Tobacco use typically begins during adolescence. There is a lack of comprehensive evidence on the use of different tobacco products among adolescents in Africa.

**Aims and Methods:** We used the most recent Global Youth Tobacco Surveys data from 53 African countries, covering 2003 – 2020, to estimate the overall and gender-specific prevalence of each type of tobacco product by country, Africa region, World Bank income group, and age group. We further used independent logit regression models to assess the determinants of the use of different types of tobacco products. The datasets covered 204,537 primary/secondary school-going adolescents aged 11 - 17 years. Pooled prevalence estimates were computed using the individual-level data.

**Results:** The overall prevalence of any tobacco use among African adolescents was 14.3% [95% CI: 13.5, 15.3]. Specifically, the prevalence for cigarette smoking was 6.4% [95% CI: 5.9, 7.0], for other smoked tobacco was 6.7% [95% CI: 6.0, 7.4], for smokeless tobacco use was 6.4% [95% CI: 5.9, 6.9], and for shisha smoking was 5.2% [95% CI: 4.4, 6.1]. The prevalence of dual use of smoked and smokeless tobacco was 3.0% [95% CI: 2.8, 3.2], and that of shisha and cigarettes was 1.5% [95% CI: 1.2, 2.0]. The prevalence of any tobacco use was higher among boys (17.4%) than girls (10.6%). Exposure to anti-tobacco smoking messages, exposure to smoking at home and school, the age restriction to tobacco purchases, and peer pressure were positively associated with the tobacco use, irrespective of product type. Being a female was a protective factor of tobacco use for almost all products. Country-level factors such as predominant religion, African region, and World Bank income group were significant factors only for cigarettes and shisha smoking.

**Conclusions:** The prevalence of tobacco use among African adolescents aged 11 - 17 years is high, but similar across different tobacco products. Peer pressure and school environment significantly influence adolescents’ decisions to participate in tobacco use. Policymakers could prioritize implementing large pictorial health warnings about tobacco dangers covering the entire packaging of different products.

**IMPLICATIONS:** African countries and regions need to strengthen tobacco control policies that are most effective in reducing tobacco consumption among young people, such as increasing tobacco taxes, textual and graphic health warnings and banning tobacco industry influence: advertising and any sponsorships. It is also important to have comprehensive surveillance systems that monitor the use of the whole range of tobacco and nicotine products overtime.

## INTRODUCTION

There are over 8 million deaths globally each year due to tobacco use and second hand tobacco smoke exposure.^1^ If no serious tobacco control efforts are taken, the number of deaths will increase to as high as 1 billion this century.^2^ Tobacco use typically starts during adolescence, and adolescents are easily addicted and ultimately exposed to long-term nicotine addiction and use.^3^ ^4^ In Africa, the tobacco industry is powerful and is increasingly targeting adolescents through the use of role models or attractive flavours for example.^2^ It is, therefore, important to monitor the use of tobacco products among young people to have the data needed to inform effective tobacco control policies.

For low- and middle-income countries, the school-based Global Youth Tobacco Survey (GYTS) provides most of the available national-level data on tobacco use among adolescents.^5^ The GYTS is a school-based survey conducted among primary/secondary school-going adolescents who are in grades that are associated with the age group 13-15 years. However, despite most GYTS datasets covering the age group 11-17 years, the most recent cross-country analyses that include datasets from African countries have focused on the 13-15 or 12-16 years age groups.^6^ ^7^ ^8^ ^9^ In addition, most of these analyses either consider the tobacco products together without disaggregated data on the different tobacco products,^6^ or consider only a few products (e.g., just cigarettes and other smoked tobacco,^7^ smokeless tobacco ^9^ or shisha (waterpipe)^8^). They also only include datasets from 11 to 38 African countries.

Our study, therefore, aims to, for the first time, provide information that covers the 11-17 years age group, disaggregated by tobacco product, and covers all African countries where at least one GYTS has been conducted. We also assess the dual use of smoked and smokeless tobacco products, as well as of cigarettes and shisha, and investigate the factors associated with the use of different tobacco products. To the best of our knowledge, this is the most comprehensive study to date to explore the prevalence of the use of tobacco products and their associated factors in Africa.

## METHODS

### Data sources

This study uses the latest GYTS data from each of the 53 African countries that have conducted at least one round of the survey.^10^ These datasets covered the period 2003 - 2020. South Sudan is the only African country that has not conducted a GYTS. The GYTS is collected through a two-stage cluster sampling design. In the first stage, schools are selected proportional to the school enrolment sizes. The second stage involves the random sampling of classes, and all students within the selected classes are eligible to participate in the self-administered questionnaire.^10^

Most of the 53 datasets comprise school-going adolescents in the age group, 11 - 17 years, with the exception of those for Carbo Verde, Chad, Ethiopia, and South Africa, which also include those outside this age group. We therefore excluded any respondents older than 17 years from the analysis for consistency across countries.

### Outcomes of interest

Our outcome variables of interest were current:

- Cigarette smoking status: this was obtained from the responses to one of the following questions: “During the past 30 days, on how many days did you smoke cigarettes?” or “During the past 30 days, did you smoke any cigarettes?”.
- Shisha smoking status: this was obtained from the responses to one of the following questions: “During the past 30 days, on how many days did you smoke shisha?” or “During the past 30 days, did you smoke shisha?”
- Use of other smoked tobacco products: this was obtained from the responses to one of the following questions: “During the past 30 days, on how many days did you smoke any smoked tobacco products other than cigarettes?” or “During the past 30 days, did you smoke any of the smoked tobacco products other than cigarettes?”
- Smokeless tobacco use status: this was obtained from the responses to one of the following questions: “During the past 30 days, on how many days did you use any smokeless tobacco products?” or “During the past 30 days, did you use any smokeless tobacco products?”
- Dual use of smoked and smokeless tobacco: individuals who had been classified as current smokers of cigarettes, shisha or other smoked tobacco products, AND current users of smokeless tobacco products were dual users of smoked and smokeless tobacco. Those who only currently used smoked products, as well as those who only currently used smokeless tobacco products were not dual users of smoked and smokeless tobacco.
- Dual use of cigarettes and shisha: individuals who had been classified as current smokers of cigarettes, AND current smokers of shisha were current dual users of cigarettes and shisha. Those who only currently used one of these products, were not dual users of cigarettes and shisha.
- Use of any tobacco product: individuals who had been classified as current users of any tobacco product, either smoked or smokeless, were classified as current users of any tobacco product. Those who did not report using any tobacco product were classified as non-users of tobacco.

For current cigarette smoking, shisha smoking, use of other smoked tobacco products and smokeless tobacco use, those whose response was one day or more to the question on the number of days they used a product in the past 30 days were classified as current smokers/users of the product, whilst those whose response was zero were current non-smokers/users. When using the question that asked whether they used a product during the past 30 days, those who answered ‘yes’ were current smokers/users of the product, whilst those who answered ‘no’ were current non-smokers/users.

Each of the outcomes of interest was defined as a dummy variable, where 1 denoted a current user and 0 a current non-user. We were also interested in heated tobacco products (HTPs) where tobacco is heated without combustion to produce aerosols containing nicotine and other chemicals, which are then inhaled by users.^11^ However, none of the surveys had data on these products.

### Independent Variables

#### Demographic variables

Guided by information from the literature on significant factors associated with tobacco use and the GYTS questionnaires, our independent variables included demographic, pro-tobacco, and anti-tobacco-use variables.^6^ ^7^ ^8^ ^9^ The demographic variables include gender, age, age squared, Africa region, and World Bank income classification. In line with de la Torre-luque et al.,^12^ we also include the predominant religion in each African country.^13^

#### Anti-tobacco use variables

We use 4 types of anti-tobacco use variables:

- Age restriction: this dummy variable was generated from the question, “During the past 30 days, did anyone refuse to sell you cigarettes because of your age?”
- Exposure to anti-tobacco media messages – a dummy variable created from the question, “During the past 30 days, did you use or hear any anti-tobacco messages on TV, radio, billboards, posters, newspapers, magazines or movies?”
- School teaching on tobacco use: this was a dummy variable generated from the question, “During the past 12 months, were you taught in any of your classes about the dangers of tobacco use?”
- Health warnings on the danger of tobacco use: this was a dummy variable generated from the question, “During the past 30 days, did you see any health warnings on cigarette/tobacco packages?”

#### Pro-tobacco use variables

We use the following 7 pro-tobacco use variables:

- Media exposure to smoking: a dummy variable generated from the question, “During the past 30 days, did you see any people using tobacco when you watched TV, videos or movies?”
- Tobacco adverts: a dummy variable generated from the question, “During the past 30 days, did you see any advertisements or promotions for tobacco products at points of sale (such as street vendors/aprons, shops, kiosks, travelling carts, etc)?”
- Offer of free tobacco products: a dummy variable generated from the question, “has a person working for the tobacco product company ever offered you a free tobacco product?”
- Public exposure to tobacco use/smoking: a dummy variable generated from the question, “during the past 7 days, on how many days has anyone smoked in your presence, at any outdoor public place such as football or basketball, fields, sidewalks, entrances to buildings, malls, parking lots, beaches, stadiums, etc?”
- School exposure to tobacco smoking: a dummy variable generated from the question, “during the past 30 days, did you see anyone smoke inside the school building or outside on school property?”
- Home exposure to tobacco smoking: a dummy variable is generated from the question, “during the past 7 days, on how many days has anyone smoked inside your home in your presence?”
- Peer/friendship smoking: a dummy variable generated from the question*, “if* one of your best friends offered you a tobacco product, would you use it?”

### Data Analysis

The GYTS datasets for the 53 countries were retrieved and homogenized in terms of key study variables, and combined into one dataset with a total sample of 204,537 school-going adolescents aged 11–17 years. We used individual-level data to compute descriptive statistics and prevalence estimates. We computed the prevalence estimates for each of the outcomes of interest for Africa, as well as by country, African region (i.e., Central, East, North, Southern, and West),^14^ World Bank income group classification^15^ as well as by gender and age group. Reported prevalence estimates include the associated 95% confidence intervals (CIs). We adjusted for the cluster variable (schools) and stratification in the variance estimation to account for the complex sampling design, and applied sample weights to account for differential probabilities of selection and participation. Data analyses were performed in Stata version 17.^16^

To assess the factors associated with tobacco use, we use data from the GYTS surveys conducted from 2013 to 2020 to ensure that we have consistent information for all variables used, including the independent variables. This is because the GYTS questionnaire changed after 2012. Given that our outcomes of interest are binary, we estimate logit regression models separately for each of the 6 products and product combinations (cigarettes smoking, other smoked tobacco use, smokeless tobacco use, shisha smoking, dual cigarettes and shisha smoking, and dual smokeless and smoked tobacco). We present the odds ratios from the Logit models. We also provide the Logit model’s marginal effects in Supplementary Table 9.

### Sensitivity analysis for prevalence estimates

Data used in this study spans the period 2003 – 2020. It is possible that, for countries with older datasets, the prevalence estimates generated might not reflect the present scenario. In order to account for this, we performed a sensitivity analysis where we excluded countries with surveys conducted before 2010.

## RESULTS

### Sample characteristics

The mean age of adolescents in the sample was 14.3 years (standard deviation = 2.2 years). About 42% of the adolescents were aged either 14 or 15 years, and there was a 50/50 gender split between boys and girls (Table 1). Close to one-third (31.89%) of the sample was from West Africa, and approximately half (48.59%) came from lower-middle-income African countries. About 20% of the sample was from surveys conducted in 2008. The highest proportion of observations was from Zimbabwe (6.3%), and the least was from Guinea-Bissau (0.7%) (Supplementary Table 1). All 53 surveys had data on cigarette smoking status, but only 16 had specific data on shisha smoking status. 41 surveys also had aggregated data on ‘other smoked tobacco products’ that are not cigarettes, and 45 surveys reported on smokeless tobacco use. Notably, no Southern African country had data on shisha smoking (Supplementary Table 2).

**Table 1:** Sample characteristics.

| Variable | Number of observations (n) | Percent (%) |
| --- | --- | --- |
| **All** | 204, 537 | 100% |
| **Gender** |  |  |
| Boys | 99,153 | 49.75% |
| Girls | 100,159 | 50.25% |
| **Age** |  |  |
| 11 years | 10,905 | 5.44% |
| 12 years | 18,526 | 9.24% |
| 13 years | 34,678 | 17.30% |
| 14 years | 42,868 | 21.39% |
| 15 years | 40,538 | 20.23% |
| 16 years | 28,566 | 14.25% |
| 17 years | 24,340 | 12.14% |
| **African Region** |  |  |
| Central Africa | 33,135 | 16.20% |
| East Africa | 40,841 | 19.97% |
| North Africa | 20,814 | 10.18% |
| Southern Africa | 44,529 | 21.77% |
| West Africa | 65,218 | 31.89% |
| **World Bank Income Group** |  |  |
| Low income | 78,520 | 38.39% |
| Lower middle | 99,384 | 48.59% |
| Upper middle | 24,148 | 11.81% |
| High income | 2,485 | 1.21% |
| **Survey Year** |  |  |
| 2003 | 5,959 | 2.94% |
| 2006 | 9,639 | 4.76% |
| 2007 | 3,965 | 1.96% |
| 2008 | 38,209 | 18.86% |
| 2009 | 16,140 | 7.96% |
| 2010 | 11,964 | 5.90% |
| 2011 | 11,837 | 5.84% |
| 2013 | 13,645 | 6.73% |
| 2014 | 20,028 | 9.88% |
| 2015 | 5,295 | 2.61% |
| 2016 | 11,896 | 5.87% |
| 2017 | 27,377 | 13.51% |
| 2018 | 10,118 | 4.99% |
| 2019 | 12,250 | 6.05% |
| 2020 | 4,320 | 2.13% |

### Prevalence of tobacco use

The overall prevalence of any tobacco use was 14.3% [95% CI: 13.5, 15.3] (Table 2). This was higher for boys (17.4% [95% CI: 16.3, 18.6]) than for girls (10.6% [95% CI: 9.5, 11.7]). The overall prevalence, as well as those for boys and girls, were highest in Southern Africa and lowest in East Africa. Upper middle-income countries had the highest, and low-income countries had the lowest overall prevalence of the use of any tobacco product. Prevalences of any tobacco use by country and gender are provided in Supplementary Table 3. The lowest prevalence was in Angola (1.2% [95% CI: 0.7, 2.0]) and the highest was in South Africa (35.2% [95% CI: 32.8, 37.8]). For boys, this ranged from 1.7% [95% CI: 0.9, 3.1] in Angola to 40.4% [95% CI: 36.9, 44.1] in South Africa. Among girls, the prevalence ranged from 0.4% [95% CI: 0.1, 1.4] in Angola to 30% [95% CI: 27.8, 32.3] in South Africa.

**Table 2:** Prevalence of use of any tobacco product by African region, World Bank income group, age group, and gender.

|  | Use of any tobacco products |  |  |
| --- | --- | --- | --- |
|  | Boys (95% CI) | Girls (95% CI) | Total (95% CI) |
| **African region** |  |  |  |
| Central Africa | 17.2% (14.7, 19.9) | 10.5% (8.8, 12.4) | 14.5% (12.5, 16.7) |
| East Africa | 10.9 (9.7, 12.2) | 6.3% (5.5, 7.2) | 8.9% (8.0, 9.9) |
| North Africa | 18.4% (15.3, 22.0) | 7.3% (4.9, 10.9) | 12.9% (10.7 to 15.5) |
| Southern Africa | 29.7% (27.7, 32.0) | 23.6% (22.1, 25.1) | 27.1% (25.6, 28.6) |
| West Africa | 15.9% (14.6, 17.3) | 10.4% (8.8, 12.1) | 13.7% (12.5, 15.0) |
| **Income Group** |  |  |  |
| High income | 24.1% (20.9, 27.5) | 13.6% (11.2, 16.3) | 18.9% (16.5, 21.5) |
| Upper middle income | 36.2% (33.4, 39.1) | 26.7% (24.9, 28.5) | 31.5% (29.6, 33.4) |
| Lower middle income | 15.1% (13.5, 16.9) | 7.8% (6.5, 9.5) | 11.8% (10.6, 13.1) |
| Low income | 14.4% (13.3, 15.5) | 8.7% (7.6, 9.8) | 12.0% (11.1, 13.0) |
| **Age Group** |  |  |  |
| 11 years | 28.9% (24.3, 34.0) | 13.7% (9.3, 19.9) | 21.1% (17.8, 25.4) |
| 12 years | 13.5% (10.5, 17.3) | 7.4% (4.9, 10.8) | 10.3% (7.9, 13.2) |
| 13 years | 12.4% (10.3, 14.9) | 7.0% (5.8, 8.4) | 9.9% (8.6%, 11.4) |
| 14 years | 14.2% (12.6, 15.9) | 10.7% (8.7, 13.1) | 12.7% (11.4, 14.1) |
| 15 years | 15.8% (14.1, 17.7) | 10.6% (9.1, 12.2) | 13.4% (12.1, 14.8) |
| 16 years | 21.2% (18.9, 23.7) | 12.4% (10.9, 14.1) | 17.3% (15.8, 19.0) |
| 17 years | 27.2% (24.9, 29.6) | 16.7% (14.7, 19.0) | 23.1% (21.4, 25.0) |
| **Overall prevalence** | **17.4% (16.3, 18.6)** | **10.6% (9.5, 11.7)** | **14.3% (13.5, 15.3)** |

Product-specific prevalence estimates by region, World Bank income group classification, and age group are also provided in Table 3. The overall prevalence of use of the different products in Africa was similar for cigarettes (6.4% [95% CI: 5.9, 7.0]), shisha (5.2% [95% CI: 4.4, 6.1]), other smoked tobacco products (6.7% [95% CI: 6.0, 7.4]), and smokeless tobacco products (6.4% [95% CI: 5.9, 6.9]). The overall prevalence of the dual use of smokeless & smoked tobacco products was 3.0% [95% CI: 2.8, 3.2], and that of the dual use of cigarettes and shisha was 1.5% [95% CI: 1.2, 2.0]. Southern Africa had the highest prevalence across all product types, except for shisha smoking where no country in that region has data. North Africa had the highest prevalence for shisha smoking. East Africa had the lowest prevalences for cigarette smoking, smokeless tobacco use and the use of other smoked tobacco products. Central Africa had the lowest prevalence of shisha smoking. Irrespective of gender, the age-specific prevalence estimates for the different tobacco products showed a similar pattern: they decreased from age 11 to 13 years, and started to increase from age 13 to 17 years (Table 3). The overall product-specific prevalence rates by country are provided in Supplementary Table 4.

**Table 3:** Prevalence of use of different tobacco products, by African region, World Bank income group, and age group.

| Location | Cigarettes (95% CI) | Smokeless (95% CI) | Other smoked (95% CI) | Shisha (95% CI) | Dual smokeless & smoked tobacco (95% CI) | Dual cigarettes & shisha (95% CI) |
| --- | --- | --- | --- | --- | --- | --- |
| **African Region** |  |  |  |  |  |  |
| Central Africa | 6.9% (5.4, 8.7) | 7.8% (6.8, 9.0) | 6.7% (5.5, 8.2) | 3.1% (2.3, 4.1) | 3.2% (2.5, 3.9) | 1.0% (0.8, 1.5) |
| East Africa | 4.2% (3.6, 4.8) | 4.1% (3.6, 4.7) | 3.4% (2.8, 4.2) | 4.5% (3.7, 5.5) | 1.5% (1.2, 1.8) | 1.5% (1.1, 1.9) |
| North Africa | 5.6% (4.3, 7.3) | 4.5% (3.4, 5.9) | 5.6% (3.9, 7.9) | 6.0% (4.6, 7.8) | 2.0% (1.7, 2.5) | 1.8% (1.2, 2.9) |
| Southern Africa | 12.7% (11.4, 14.0) | 12.8% (12.0, 13.7) | 14.0% (13.0, 15.0) | X | 7.3% (6.6, 8.0) | X |
| West Africa | 5.5% (4.9, 6.1) | 6.3% (5.5, 7.16) | 6.2% (5.5, 6.9) | 4.7% (3.6, 6.1) | 2.8% (2.4, 3.3) | 1.0% (0.7, 1.4) |
| **Income Group** |  |  |  |  |  |  |
| High income | 15.6% (13.4, 18.2) | 1.9% (1.3, 2.6) | 9.8% (8.2, 11.7) | 13.6% (11.8, 15.6) | 1.3% (0.9, 2.0) | 5.9% (4.8, 7.2) |
| Upper middle income | 15.0% (13.4, 16.7) | 14.3% (13.3, 15.3) | 17.6% (16.2, 19.1) | 4.7% (3.7, 5.9) | 8.4% (7.6, 9.4) | 1.8% (1.3, 2.6) |
| Lower middle income | 5.1% (4.4, 6.0) | 4.8% (4.2, 5.5) | 4.9% (4.1, 6.0) | 5.6% (4.6, 6.8) | 2.1% (1.8, 2.3) | 1.6% (1.1, 2.2) |
| Low income | 5.3% (4.7, 6.0) | 6.0% (5.4, 6.8) | 5.8% (5.0, 6.6) | 3.9% (3.0, 5.0) | 2.4% (2.0, 2.8) | 1.2% (0.9, 1.7) |
| **Age Group** |  |  |  |  |  |  |
| 11 years | 7.3% (5.4, 9.8) | 12.6% (9.3, 16.9) | 11.3% (8.3, 15.4) | 9.3% (6.6, 12.9) | 7.0% (4.7, 10.5) | 2.2% (0.7, 6.1) |
| 12 years | 3.4% (2.6, 4.5) | 5.0% (3.8, 6.7) | 5.3% (3.6, 7.7) | 3.3% (2.0, 5.6) | 2.1% (1.5, 2.8) | 1.0% (0.5, 2.1) |
| 13 years | 3.7% (3.2, 4.3) | 3.7% (3.1, 4.5) | 4.7% (3.4, 6.3) | 3.7% (2.8, 4.9) | 1.4% (1.5, 2.8) | 0.5% (0.4, 0.8) |
| 14 years | 5.0% (4.4, 5.8) | 5.5% (4.5, 6.8) | 5.5% (4.7, 6.5) | 4.8% (3.9, 5.8) | 2.2% (1.9, 2.6) | 1.1% (0.8, 1.6) |
| 15 years | 6.5% (5.4, 7.6) | 5.7% (5.0, 6.4) | 6.0% (5.2, 6.9) | 6.0% (4.5, 8.1) | 2.8% (2.3, 3.3) | 2.0% (1.2, 3.2) |
| 16 years | 8.0% (7.0% 9.1) | 8.2% (7.1, 9.5) | 8.1% (7.2, 9.0) | 5.8% (4.6, 7.3) | 3.8% (3.2, 4.4) | 2.5% (1.6, 3.7) |
| 17 years | 14.0% (12.6, 15.5) | 10.0% (9.0, 11.0) | 11.6% (10.3, 13.0) | 6.5% (5.0, 8.3) | 6.1% (5.4, 6.9) | 2.8% (1.8, 4.2) |
| **Overall Prevalence** | **6.4% (5.9, 7.0)** | **6.4% (5.9, 6.9)** | **6.7% (6.0, 7.4)** | **5.2% (4.4, 6.1)** | **3.0% (2.8, 3.2)** | **1.5% (1.2, 2.0)** |

Notes: X indicates that data was not available.

**Table 4:** Factors associated with cigarette smoking, other smoked and smokeless tobacco use.

|  | Cigarettes | Other Smoked Tobacco | Smokeless Tobacco |
| --- | --- | --- | --- |
| VARIABLES | Odds Ratio<br>(95% CI) | Odds Ratio<br>(95% CI) | Odds Ratio<br>(95% CI) |
| Age | 0.4<br>(0.1, 1.4) | 0.3<br>(0.1, 1.3) | 0.7<br>(0.1, 5.1) |
| Age Squared | 1.0<br>(0.9, 1.1) | 1.0<br>(1.0, 1.1) | 1.0<br>(1.0, 1.1) |
| Female (Baseline: Male) | 0.3*<br>(0.2, 0.3) | 0.4*<br>(0.3, 0.6) | 0.9<br>(0.6, 1.3) |
| *Region (Base: Central Africa)* |  |  |  |
| East Africa | 0.4*<br>(0.3, 0.7) | 0.8<br>(0.5, 1.3) | 0.7*<br>(0.5, 1.0) |
| North Africa | 0.6<br>(0.3, 1.1) | 1.1<br>(0.6, 2.0) | 0.9<br>(0.6, 1.4) |
| Southern Africa | 0.5*<br>(0.3, 0.9) | 1.1<br>(0.7, 1.7) | 0.9<br>(0.6, 1.3) |
| West Africa | 0.5*<br>(0.3, 1.0) | 1.3<br>(0.9, 1.8) | 0.7<br>(0.4, 1.2) |
| *World Bank Income Group (Base: Low income)* |  |  |  |
| Lower middle income | 0.5*<br>(0.4, 0.7) | 0.9<br>(0.7, 1.3) | 0.9<br>(0.7, 1.3) |
| Upper middle income | 1.4<br>(0.9, 2.0) | 1.5<br>(1.0, 2.4) | 0.7<br>(0.4, 1.1) |
| Christian (Base: Muslim & Hindu) | 0.8<br>(0.6, 1.2) | 0.8*<br>(0.6, 1.0) | 1.0<br>(0.7, 1.5) |
| Age Restriction | 4.7*<br>(3.6, 6.2) | 2.4*<br>(1.6, 3.5) | 2.4*<br>(1.7, 3.5) |
| Anti-tobacco use messages | 0.7*<br>(0.6, 0.8) | 1.183<br>(0.9, 1.5) | 1.0<br>(0.7, 1.3) |
| School teaching on tobacco use | 0.9<br>(0.7, 1.1) | 1.0<br>(0.7, 1.5) | 1.2<br>(0.8, 1.6) |
| Health warnings | 2.0*<br>(1.5, 2.7) | 1.5*<br>(1.2, 1.9) | 1.8*<br>(1.3, 2.4) |
| Media exposure to smoking | 1.1<br>(0.9, 1.3) | 0.9<br>(0.6, 1.2) | 0.8<br>(0.6, 1.1) |
| Tobacco adverts | 0.9<br>(0.7, 1.2) | 1.2<br>(0.8, 1.8) | 1.1<br>(0.8, 1.6) |
| Free tobacco product offer | 1.8*<br>(1.3, 2.5) | 3.7*<br>(2.6, 5.3) | 4.6*<br>(3.3, 6.4) |
| Public exposure to smoking | 2.2*<br>(1.7, 2.9) | 0.9<br>(0.6, 1.5) | 1.1<br>(0.7, 1.6) |
| School exposure to smoking | 2.0*<br>(1.5, 2.7) | 1.6*<br>(1.1, 2.3) | 1.7*<br>(1.3, 2.3) |
| Home exposure to smoking | 2.5*<br>(1.8, 3.4) | 1.9*<br>(1.2, 2.8) | 1.2<br>(0.9, 1.7) |
| Peer smoking | 8.7*<br>(6.9, 11.0) | 2.7*<br>(2.1, 3.4) | 3.0*<br>(2.3, 3.9) |
| Observations | 82,424 | 81,272 | 83,649 |

Notes: The 95% Confidence Intervals (CI) of the Odds Ratios are in parentheses. \* indicates statistical significance, that is, p-value < 0.05.

Figure 1 presents the regional prevalence estimates for the use of different tobacco products by gender. The prevalence of cigarette smoking was highest among boys in all regions. The prevalence of smokeless tobacco use was most common among girls in 4 regions except Southern Africa. The prevalence rates among girls in Southern Africa were higher than that of boys in other regions for all products. Gender-specific country-level estimates for different tobacco products are provided in Supplementary Tables 5 for boys and 6 for girls. For Africa as a whole, tobacco use prevalence was higher among boys than for girls across all product types. Among boys, cigarette smoking was more common (9.2%), followed by other smoked tobacco (8.3%), smokeless (7.3%), and shisha (6.5%), in that order. Among girls, the prevalence of smokeless tobacco was more common (5.1%), followed by other smoked (4.8%), shisha (3.6%), and cigarettes (3.4%), in that order.

**Figure 1:**
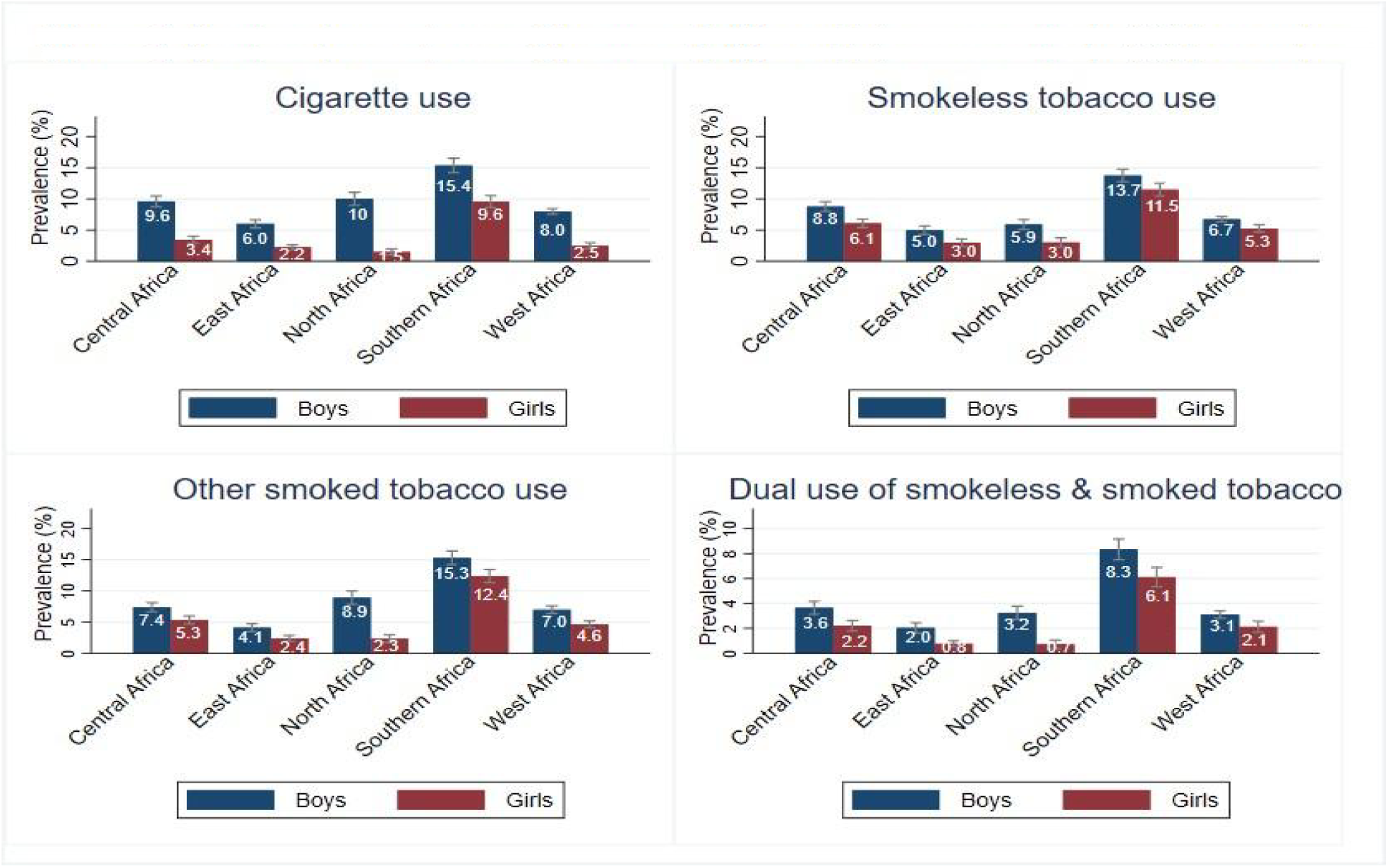
Regional prevalence of the use of different tobacco products (%) by gender

### Factors associated with the use of tobacco products

#### Cigarette smoking

For cigarettes, girls were significantly less likely to smoke than boys (odds ratio (OR) 0.3, [95% CI: 0.2, 0.3]). Adolescents from East Africa (OR 0.4, [95% CI = 0.3, 0.7]), Southern Africa (OR 0.5, [95% CI=0.3, 0.9]), and West Africa (OR 0.5, [95% CI=0.3, 1.0]) were less likely to smoke than those from Central Africa. There was no statistically significant difference between North Africa and Central Africa in the likelihood of an adolescent being a cigarette smoker. Adolescents from lower-middle-income African countries were less likely to smoke (OR 0.5, [95% CI = 0.4, 0.7]) than those from low-income countries. There was no statistically significant difference between upper-middle and lower-middle-income countries on the likelihood of an adolescent being a current cigarette smoker.

Adolescents who were refused the purchase of cigarettes because of their age (aged below 18) (either for themselves or on behalf of other people) were more likely to smoke cigarettes (OR 4.7, [95% CI = 3.6, 6.2]) than those who were not. Those who were exposed to anti-tobacco smoking messages were less likely to smoke (OR 0.8, [95% CI = 0.6, 0.8]) than those who were not. Adolescents who had seen the health warnings about the dangers of tobacco use were more likely to use cigarettes (OR 2.0, 95% CI= 1.5, 2.7]) than those who had not.

When considering the pro-tobacco use variables, adolescents who had been offered free tobacco products were more likely to smoke (OR 1.8, [95% CI= 1.3, 2.5]) than those who had not. Exposure to tobacco use in public places (OR 2.2, [95% CI= (1.7, 2.9)]), at school (OR 2.0, [95% CI: 1.5, 2.7]), and at home (OR 2.5, [95% CI: 1.8, 3.4]) had higher of odds of cigarette smoking. Having smoking friends/peers was associated with higher odds of current smoking among adolescents (OR 8.685, [95 % CI: 6.879, 10.97]) compared to not having any smoking friends.

#### Shisha smoking

Girls were less likely to smoke shisha (OR 0.7, [95% CI: 0.5, 0.9]) than boys. Adolescents from North Africa (OR 3.2, [95% CI: 1.7, 5.8]) and West Africa (OR 2.1, [95% CI=1.3, 3.5]) were more likely to smoke than those from Central Africa. The association was statistically insignificant between residing in East Africa and shisha smoking. Adolescents from lower-middle-income countries were more likely to use shisha (OR 1.4, [95% CI = 1.1, 2.0]) than those from low-income countries. Students from countries dominated by Christianity were more likely to consume shisha (OR 1.7, [95% CI: 1.2, 2.3]) than those dominated by Islam and Hinduism.

Anti-tobacco use factors indicate that adolescents who were refused to buy cigarettes because of their age (aged below 18) (either for themselves or on behalf of other people) were more likely to smoke shisha (OR 5.2, [95% CI = 3.4, 7.7]) than those who were not. Those who were taught about tobacco dangers at school were less likely to use shisha (OR 0.6, [95% CI = 0.5, 0.8]) than those who were not. Adolescents who had seen the health warnings about the dangers of tobacco use were more likely to use shisha (OR 1.7, [95% CI= 1.1, 2.7]) than those who had not.

In terms of pro-tobacco use variables, those who had been offered free tobacco products were more likely to consume shisha (OR 2.9, [95% CI= 2.0, 4.2]) than those who had not. Exposure to tobacco use in public places (OR 1.4, [95% CI= (1.0, 2.0)]), at school (OR 1.3, [95% CI: 1.0, 1.7]), and at home (OR 2.0, [95% CI: 1.5, 2.6]) had a significant positive association with shisha consumption. Having smoking friends/peers was associated with higher odds of current smoking among adolescents (OR 3.7, [95 % CI: 2.5, 5.4]) compared to not having any smoking friends.

#### Other smoked tobacco products

For other smoked tobacco products, girls were less likely to smoke them (OR 0.4, [95% CI = 0.3, 0.6]) than boys. Adolescents from countries that predominantly have Christians were less likely to smoke (OR 0.8, 95% CI = [0.6, 1.0]) compared to those that predominantly have Muslims and Hindus. Those who were refused the purchase of cigarettes because of their age were more likely to smoke other smoked products (OR 2.4, [95% CI = 1.6, 3.5]) than those who were not. Those who had seen health warnings about the dangers of tobacco were more likely to smoke (OR 1.5, [95% CI: 1.2, 1.9]) than those who had not.

Regarding pro-tobacco variables, adolescents who had been offered free tobacco products were more likely to use other smoked tobacco (OR 3.7 [95% CI= 2.6, 5.3) than those who had not. Those who had been exposed to tobacco use at school (OR 1.6, [95% CI: 1.1, 2.3]) and at home (OR 1.9, [95% CI: 1.2, 2.8]) were more likely to use other smoked tobacco than those who had not been exposed to smoking in any of these places. Having smoking peers was associated with higher odds of other smoked tobacco use (OR 2.7, [95 % CI: 2.1, 3.4]) compared to not having any smoking friends.

#### Smokeless tobacco use

For smokeless tobacco products, adolescents who reside in East Africa (OR 0.7, [95% CI: 0.5, 1.0]) were less likely to use smokeless tobacco than those in Central Africa. Residing in other African regions (North, West, and South) was not significantly associated with smokeless tobacco use. Only two anti-tobacco use variables were significantly associated with consuming smokeless tobacco. Those who were restricted from purchasing cigarettes because of their age were more likely to use smokeless tobacco than those who were not. Those who had seen health warnings about the dangers of tobacco were more likely to consume smokeless tobacco (OR 1.8, [95% CI: 1.3, 2.4]) than those who had not. Concerning the pro-tobacco variables, those who had been offered free tobacco products were likelier to use smokeless tobacco (OR 4.6, [95% CI= 3.3, 6.4]) than those who had not. Being exposed to tobacco use at school was associated with higher odds of using smokeless tobacco (OR 4.6, [95% CI= 3.3, 6.4]) compared to not being exposed.

#### Dual use

For the factors associated with the dual use of tobacco products, dual smoked and smokeless, and dual shisha and cigarettes, the same anti-tobacco and pro-tobacco factors that were significantly associated with the use of single products were associated with the dual use (see Table 5). Seeing tobacco adverts on any media platform within a month had a significant positive association only with dual cigarette and shisha smoking (OR 2.2, [95% CI = 1.1, 4.3]) but a statistically insignificant association with other products. Being taught about tobacco dangers at school had a significant negative association with dual cigarettes and shisha smoking (OR 0.6, [95% CI: 0.1, 1.0]) but an insignificant association for other products.

**Table 5:**
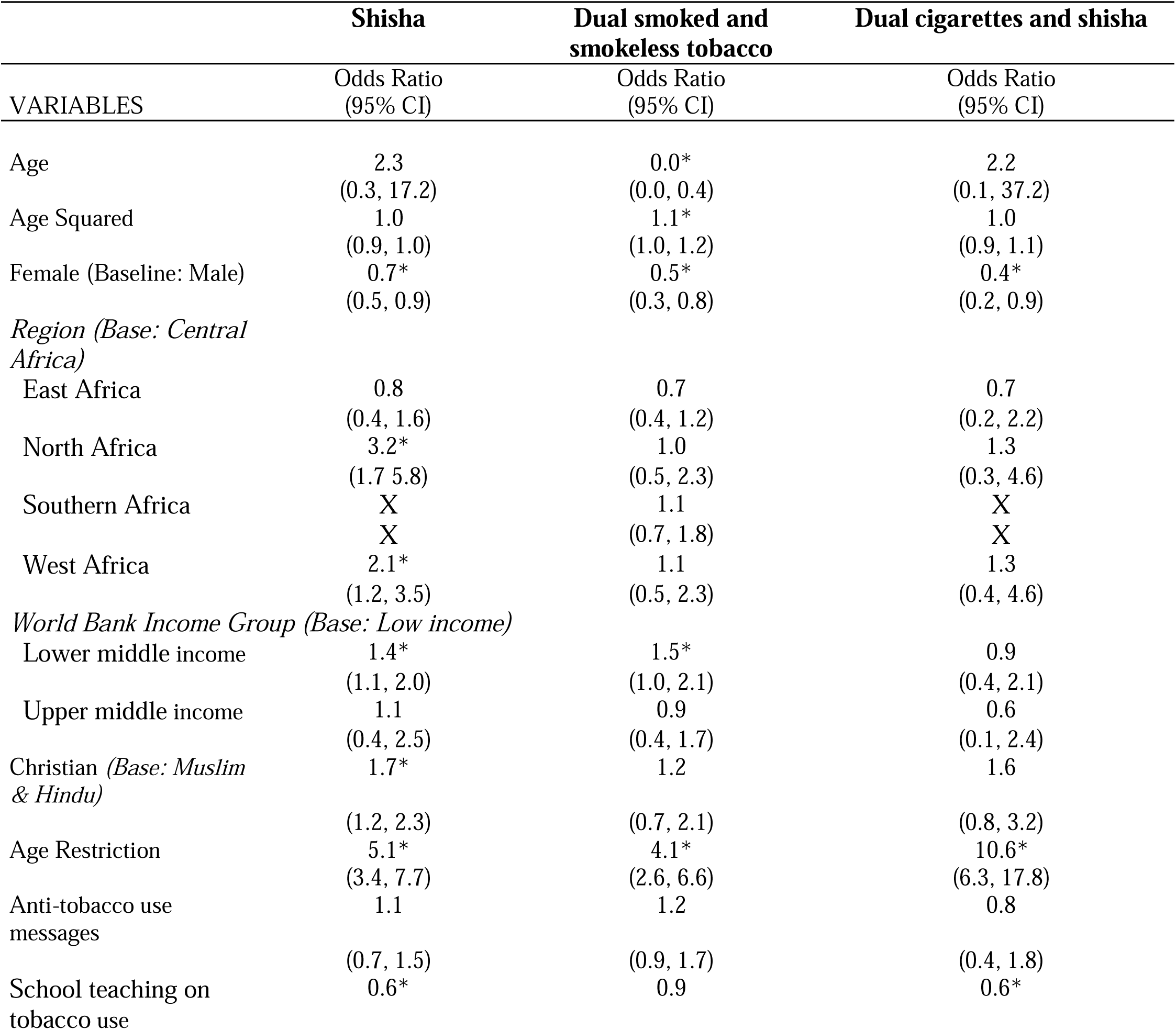
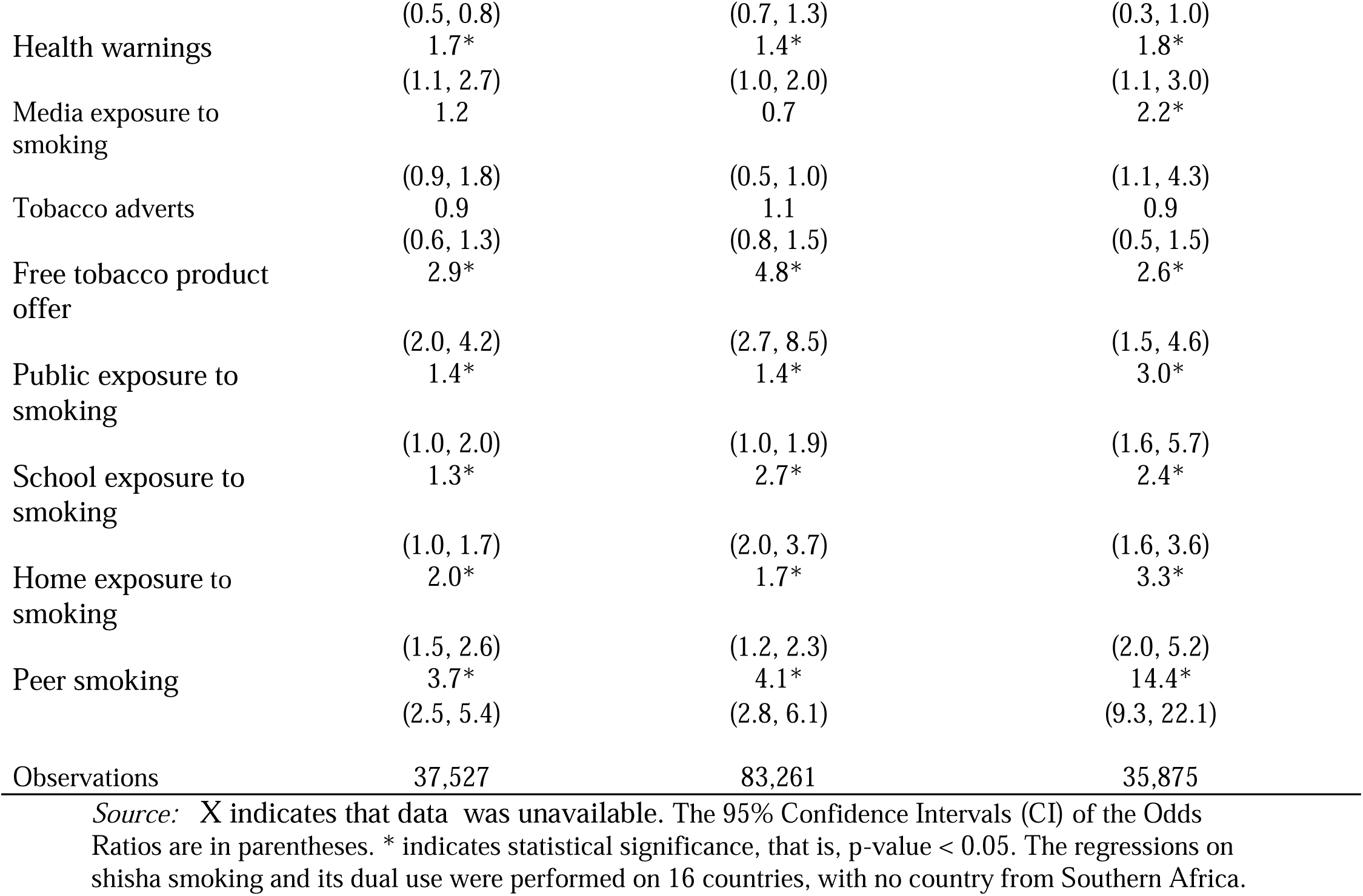
Factors associated with shisha smoking, dual smoked and smokeless tobacco use, and dual cigarette and shisha smoking.

### Sensitivity analysis results

The prevalence estimates generated with the full dataset were similar to those generated when datasets collected before 2010 were excluded. In the instance that there were differences in the prevalence estimates, the difference was not more than 2 percentage points. Thus, overall, our results are robust to excluding the data that is at least 10 years old.

## DISCUSSION

We found a prevalence of current use of any tobacco of 14.3% [95% CI: 13.5, 15.3]. For cigarettes, the prevalence of current use was 6.4% [95% CI: 5.9, 7.0], and this was 5.2% [95% CI: 4.4, 6.1] for shisha, 6.7% [95% CI: 6.0, 7.4] for other smoked tobacco products, and 6.4% [95% CI: 5.9, 6.9] for smokeless tobacco products. The prevalence of tobacco use, including for many of the individual tobacco product types, was highest in Southern Africa and lowest in East Africa, except for shisha, where it was highest in North Africa and lowest in Central Africa. Tobacco use prevalence was higher for boys than for girls, both overall and across all product types. The age-specific prevalence estimates for the different tobacco products decreased from age 11 to 13 years, and then increased from age 13 to 17 years.

Our overall prevalence rates for Africa, including the product-, gender- and age-specific prevalence rates, align with those from other studies. For example, another study reported Africa-specific prevalence rates of 16.2% for any tobacco use, 10.7% for other smoked products, and 8.6% for cigarettes among boys; and 10.3 for any tobacco use, 7.9% for other smoked products, and 3.7% for cigarettes among girls.^7^ Other studies have also reported a prevalence of smokeless tobacco use of 4.1% and 5.1% in Africa for the 13-15^6^ and 12-16 years^9^ age groups, respectively. Another analysis of data from 22 African countries reported cigarette smoking and any tobacco use prevalence estimates that are higher than those obtained in the current study (10.9% and 19.1%, respectively).^6^ The difference in these prevalence rates is because our study includes 31 more African countries that were not included in this study.

In our study, we explored the dual use of tobacco products among adolescents in Africa for the first time and found a prevalence of 3.0% for the dual use of smoked and smokeless tobacco products. Additionally, we found a prevalence of 1.5% for the dual use of cigarettes and shisha. The availability and heavy marketing of more tobacco and nicotine products could result in a continued increase in dual tobacco use among the youth,^17^ and result in high exposure to addiction and lower chances of quitting among young people in Africa.^18^ In addition, tobacco use and alcohol consumption in adolescents often co-occur, and both are strongly and positively associated with the use of other psychoactive substances such as marijuana.^19^ Understanding this phenomenon, including the interplay among these products in terms of initiation and continued use, will be important for effective tobacco control efforts. For example, there might be a need for the development of tobacco cessation programmes that account for the dual use of tobacco products and their co-consumption with other psychoactive substances.

Our study confirms findings from other African studies that girls are currently less likely to use smoked tobacco products than males.^6^ ^20^ However, this might change in the future because of changing social and cultural norms. There is also an emergence of tobacco products that are viewed as more acceptable for women to use. For example, although we did not find this in our current study, some studies in Nigeria have shown that among the youths, females are as likely to be current shisha smokers as males ^21^ and that shisha use among females is perceived as more accepted by society, and more comfortable to females when compared to cigarette smoking.^22^ In addition, with the plateauing or decline of tobacco use in many high-income countries, there are indications that the tobacco industry is now aggressively marketing to women and girls in low and middle-income countries.^23^ Our analysis showed that for some countries, the gap between boys and girls in terms of overall tobacco use prevalence has closed: for example, Zimbabwe with a prevalence of 22.6% [95% CI: 17.4, 28.8] among boys and 16.9% [95% CI: 12.9, 21.8] among girls.

In our study, those who had been denied the purchase of cigarettes because of their age were more likely to be current tobacco users than those who had not, which contradicts evidence from another study focusing on South Asia, where age restriction was found to be protective of adolescents from smoking.^24^ Second, the results for the health warnings also suggested that seeing health warnings about tobacco dangers is a positive predictor of tobacco use across all products. For policy, this implies tobacco control policymakers should consider textual and graphic health warnings, as they are considered one of the effective tools to reduce tobacco use.^25^ The unexpected results of health warnings and age restrictions may be because of how the GYTS questionnaire was designed. It is plausible that this question was asked even to those who did not smoke.

School teaching about the dangers of tobacco use had a negative association with tobacco use, especially shisha smoking and dual shisha and cigarette smoking. This aligns with evidence from countries like Lesotho,^26^ and cross-country findings from 73 countries,^8^ 4 South Asian countries,^24^ and 138 countries.^9^ In fact, our results about school pro- and anti-tobacco use factors suggest that the school environment plays a role in reducing the odds of cigarette smoking and dual shisha and cigarette smoking.

In contrast, exposure to anyone smoking at home and peer smoking have strong positive associations with tobacco use, irrespective of tobacco product type. Public exposure to smoking was also a statistically highly significant positive predictor of specific tobacco products: cigarette smoking, shisha smoking, and the dual use of smoked and smokeless tobacco. This supports existing evidence on the importance of the home and social environment in shaping adolescents’ tobacco use behaviours.^6^ ^8^ ^20^ As with other studies,^9^ our results also show that tobacco industry interference through the offer of free tobacco products can significantly increase the likelihood of use across all tobacco product types. This is critical for African tobacco control policy to fight tobacco industry influence since the industry targets African adolescents to retain them as their life time consumers.^2^

In Africa, the age of initiation among young people ranges from as low as 7 years to about 16 years.^27^ ^28^ ^29^ African countries and regions need to strengthen their tobacco control responses to curb tobacco initiation and use among young people and to reduce tobacco use prevalence. It is also essential to have comprehensive surveillance systems that monitor the use of the whole range of tobacco and nicotine products overtime. Data has to cover all adolescents (10 - 19 year olds) not only those that are 13 - 15 years old. Data that includes adolescents who are out of school is also needed: some studies suggest that they might be more vulnerable to tobacco use than those who are in school.^30^ ^31^

The analysis from our study covers 11 - 17-year-old primary/secondary school-going adolescents in grades associated with age groups 13 - 15 years. In addition, the datasets used for some countries are old and are from different years. This means that the results might not be generalizable to all the adolescents at the country or regional levels, particularly in the context of high out-of-school rates; and might not reflect the current use situation accurately. Nonetheless, our study provides comprehensive estimates, using currently available data, for different types of tobacco products in Africa, and contributes to the evidence needed for tobacco control policy decisions in Africa.

## CONCLUSION

The prevalence of tobacco use among African adolescents is high and similar for different tobacco products. Some demographic factors only matter for using specific products, while anti-tobacco factors, although counterintuitive, matter for tobacco use irrespective of product type. Peer influence, school and home environment, and health warnings about tobacco use are risk factors for using tobacco, irrespective of product type among adolescents. Policymakers should develop pictorial and graphical health warnings about tobacco dangers to discourage tobacco use and educate the entire adolescent peer group.

## DATA AVAILABILITY

All the data are publicly available on the WHO Microdata repository and can be accessed here.

## ACKNOWLEDGMENTS

This study was conducted as part of the Data on Youth and Tobacco in Africa (DaYTA) program. The DaYTA program seeks to empower stakeholders to make timely, data-driven decisions by using evidence to inform policy and advance tobacco control efforts in sub-Saharan Africa. The program achieves this by addressing data gaps related to tobacco use among 10- to 17-year-olds in Kenya, Nigeria, and the Democratic Republic of Congo. The DaYTA program is led by Development Gateway: an IREX Venture (DG), a global non-profit organization that specializes in data for development.

