## Supplementary Information for "Prevalence and determinants of tobacco use among school-going adolescents in 53 African countries: evidence from Global Youth Tobacco Surveys"

**Supplementary Table 1: Sample distribution by country**

| Variable | Number of observations (n) | Percent (%) |
| --- | --- | --- |
| **All** | 204, 537 | 100% |
| **Country (GYTS Year)** |  |  |
| Algeria (2013) | 6228 | 3.04% |
| Angola (2010) | 1427 | 0.70% |
| Benin (2003) | 4329 | 2.12% |
| Botswana (2008) | 2207 | 1.08% |
| Burkina Faso (2009) | 3656 | 1.79% |
| Burundi (2008) | 2521 | 1.23% |
| Cameroon (2014) | 2922 | 1.43% |
| Carbo Verde (2007) | 1967 | 0.96% |
| Central African Republic [CAR] (2008) | 2027 | 0.99% |
| Chad (2019) | 1937 | 0.95% |
| Comoros (2015) | 2810 | 1.37% |
| Congo (2019) | 6396 | 3.13% |
| Cote D’Ivoire (2009) | 3619 | 1.77% |
| Democratic Republic of Congo [DRC] (2008) | 3890 | 1.90% |
| Djibouti (2013) | 1818 | 0.89% |
| Egypt (2014) | 2471 | 1.21% |
| Equatorial Guinea (2008) | 3136 | 1.53% |
| Eritrea (2006) | 9639 | 4.71% |
| Eswatini (2009) | 2286 | 1.12% |
| Ethiopia (2003) | 1630 | 0.80% |
| Gabon (2014) | 1781 | 0.87% |
| Gambia (2017) | 12585 | 6.15% |
| Ghana (2017) | 5664 | 2.77% |
| Guinea (2008) | 3648 | 1.78% |
| Guinea-Bissau (2008) | 1390 | 0.68% |
| Kenya (2013) | 1895 | 0.93% |
| Lesotho (2008) | 3426 | 1.68% |
| Liberia (2008) | 1739 | 0.85% |
| Libya (2010) | 2012 | 0.98% |
| Madagascar (2018) | 2920 | 1.43% |
| Malawi (2009) | 2482 | 1.21% |
| Mali (2008) | 4071 | 1.99% |
| Mauritania (2018) | 3740 | 1.83% |
| Mauritius (2016) | 4141 | 2.02% |
| Morocco (2016) | 3915 | 1.91% |
| Mozambique (2013) | 5599 | 2.74% |
| Namibia (2008) | 2411 | 1.18% |
| Niger (2008) | 2174 | 1.06% |
| Nigeria (2009) | 5459 | 2.67% |
| Rwanda (2008) | 2284 | 1.12% |
| Sao Tome and Principe (2008) | 8525 | 4.17% |
| Senegal (2020) | 4320 | 2.11% |
| Seychelles (2015) | 2485 | 1.21% |
| Sierra Leone (2017) | 6680 | 3.27% |
| Somalia (2007) | 1998 | 0.98% |
| South Africa (2011) | 8460 | 4.14% |
| Sudan (2009) | 1923 | 0.94% |
| Tanzania (2016) | 3840 | 1.88% |
| Togo (2019) | 3917 | 1.92% |
| Tunisia (2017) | 2448 | 1.20% |
| Uganda (2018) | 3458 | 1.69% |
| Zambia (2011) | 3377 | 1.65% |
| Zimbabwe (2014) | 12854 | 6.28% |

**Supplementary Table 2: Recent data availability of different tobacco products in African GYTS, 2003 - 2020**

| **Location** | **Cigarettes** | **Smokeless tobacco** | **Other smoked tobacco** | **Shisha** |
| --- | --- | --- | --- | --- |
| **Central Africa** |  |  |  |  |
| Burundi (2008) | ✔ | 🞩 | 🞩 | 🞩 |
| Cameroon (2014) | ✔ | ✔ | ✔ | 🞩 |
| CAR (2008) | ✔ | ✔ | ✔ | 🞩 |
| Chad (2019) | ✔ | ✔ | ✔ | ✔ |
| Congo (2019) | ✔ | ✔ | ✔ | ✔ |
| DRC (2008) | ✔ | ✔ | ✔ | 🞩 |
| Equatorial Guinea (2008) | ✔ | ✔ | 🞩 | 🞩 |
| Gabon (2014) | ✔ | ✔ | ✔ | ✔ |
| Sao Tome and Principe (2008) | ✔ | ✔ | ✔ | 🞩 |
| **Total datasets in the region** | **9** | **8** | **7** | **3** |
| **East Africa** |  |  |  |  |
| Comoros (2015) | ✔ | ✔ | 🞩 | 🞩 |
| Djibouti (2013) | ✔ | ✔ | ✔ | ✔ |
| Eritrea (2006) | ✔ | 🞩 | 🞩 | 🞩 |
| Ethiopia (2003) | ✔ | 🞩 | 🞩 | 🞩 |
| Kenya (2013) | ✔ | ✔ | ✔ | ✔ |
| Madagascar (2018) | ✔ | ✔ | ✔ | 🞩 |
| Mauritius (2016) | ✔ | ✔ | ✔ | 🞩 |
| Rwanda (2008) | ✔ | ✔ | ✔ | 🞩 |
| Seychelles (2015) | ✔ | ✔ | ✔ | ✔ |
| Somalia (2007) | ✔ | 🞩 | 🞩 | 🞩 |
| Sudan (2009) | ✔ | ✔ | 🞩 | ✔ |
| Tanzania (2016) | ✔ | ✔ | ✔ | 🞩 |
| Uganda (2018) | ✔ | ✔ | ✔ | ✔ |
| **Total datasets in the region** | **13** | **10** | **8** | **5** |
| **North Africa** |  |  |  |  |
| Algeria (2013) | ✔ | ✔ | ✔ | 🞩 |
| Egypt (2014) | ✔ | ✔ | ✔ | ✔ |
| Libya (2010) | ✔ | ✔ | 🞩 | ✔ |
| Mauritania (2018) | ✔ | ✔ | ✔ | 🞩 |
| Morocco (2016) | ✔ | ✔ | ✔ | ✔ |
| Tunisia (2017) | ✔ | ✔ | ✔ | ✔ |
| **Total datasets in the region** | **6** | **6** | **5** | **4** |
| **Southern Africa** |  |  |  |  |
| Angola (2010) | ✔ | 🞩 | 🞩 | 🞩 |
| Botswana (2008) | ✔ | ✔ | ✔ | 🞩 |
| Eswatini (2009) | ✔ | ✔ | ✔ | 🞩 |
| Lesotho (2008) | ✔ | ✔ | ✔ | 🞩 |
| Malawi (2009) | ✔ | ✔ | ✔ | 🞩 |
| Mozambique (2013) | ✔ | ✔ | ✔ | 🞩 |
| Namibia (2008) | ✔ | ✔ | ✔ | 🞩 |
| South Africa (2011) | ✔ | ✔ | ✔ | 🞩 |
| Zambia (2011) | ✔ | ✔ | ✔ | 🞩 |
| Zimbabwe (2014) | ✔ | ✔ | ✔ | 🞩 |
| **Total datasets in the region** | **10** | **9** | **9** | **0** |
| **West Africa** |  |  |  |  |
| Benin (2003) | ✔ | 🞩 | 🞩 | 🞩 |
| Burkina Faso (2009) | ✔ | ✔ | ✔ | 🞩 |
| Carbo Verde (2007) | ✔ | 🞩 | 🞩 | 🞩 |
| Cote D’Ivoire (2009) | ✔ | ✔ | ✔ | 🞩 |
| Gambia (2017) | ✔ | ✔ | ✔ | 🞩 |
| Ghana (2017) | ✔ | ✔ | ✔ | ✔ |
| Guinea (2008) | ✔ | ✔ | ✔ | 🞩 |
| Guinea-Bissau (2008) | ✔ | ✔ | ✔ | 🞩 |
| Liberia (2008) | ✔ | ✔ | ✔ | 🞩 |
| Mali (2008) | ✔ | ✔ | ✔ | 🞩 |
| Niger (2008) | ✔ | 🞩 | 🞩 | 🞩 |
| Nigeria (2009) | ✔ | ✔ | ✔ | 🞩 |
| Senegal (2020) | ✔ | ✔ | ✔ | ✔ |
| Sierra Leone (2017) | ✔ | ✔ | ✔ | ✔ |
| Togo (2019) | ✔ | ✔ | ✔ | ✔ |
| **Total datasets in the region** | **15** | **12** | **12** | **4** |
| **Total available datasets in Africa** | **53** | **45** | **41** | **16** |

*Notes:* ✔ indicates that data is available, and 🞩 indicates that data is not available

**Supplementary** **Table 3: Prevalence of use of any tobacco product, by country and African region**

|  | **Use of any tobacco products** | | | |
| --- | --- | --- | --- | --- |
|  | **Boys (95% CI)** |  | **Girls (95% CI)** | **Total (95% CI)** |
| **Central Africa** |  |  |  |  |
| Burundi (2008) | 8.5% (4.9, 14.5) |  | 2.7% (1.5, 5.0) | 5.9% (3.6, 9.3) |
| Cameroon (2014) | 13.6% (10.0, 18.2) |  | 5.9% (4.5, 7.7) | 10.0% (7.5, 13.4) |
| CAR (2008) | 28.2% (23.0, 33.9) |  | 25.5% (12.9, 44.3) | 27.3% (19.5, 36.8) |
| Chad (2019) | 10.60% (10.6, 10.6) |  | 12.2% (12.2, 12.2) | 11.7% (11.7, 11.7) |
| Congo (2019) | 14.0% (11.2, 17.3) |  | 11.1% (8.9, 13.7) | 13.4% (11.0, 16.3) |
| DRC (2008) | 36.7% (31.0, 42.8) |  | 27.6% (22.5, 33.5) | 32.7% (28.2, 37.4) |
| Equatorial Guinea (2008) | 33.3% (26.9, 40.3) |  | 23.7% (18.2, 30.3) | 29.1% (23.7, 35.2) |
| Gabon (2014) | 8.8% (6.7, 11.5) |  | 6.9% (5.2, 9.3) | 8.1% (6.4, 10.1) |
| Sao Tome and Principe (2008) | 33.9 (33.9, 33.9) |  | 25.3% (25.3, 25.3) | 29.1% (29.1, 29.1) |
| **Regional Prevalence** | **17.2% (14.7, 19.9)** |  | **10.5% (8.8, 12.4)** | **14.5% (12.5, 16.7)** |
| **East Africa** |  |  |  |  |
| Comoros (2015) | 19.9% (16.7, 23.7) |  | 12.8% (10.1, 16.0) | 16.4% (13.5,19.7) |
| Djibouti (2013) | 20.8% (17.7, 24.4) |  | 18.9% (14.9, 23.6) | 20.4% (17.6, 23.6) |
| Eritrea (2006) | 2.6% (2.0, 3.5) |  | 0.7% (0.3, 1.5) | 2.0% (1.4, 2.7) |
| Ethiopia (2003) | 3.5% (2.4, 5.2) |  | 1.1% (0.5, 2.3) | 2.6% (1.7, 3.9) |
| Kenya (2013) | 11.1 % (8.9, 13.8) |  | 8.7% (6.9, 10.9) | 10.1% (8.2, 12.3) |
| Madagascar (2018) | 20.2% (16.7, 24.3) |  | 6.2% (4.7, 8.1) | 13.2% (11.1, 15.7) |
| Mauritius (2016) | 27.9% (22.3, 34.2) |  | 10.7 (7.3, 15.3) | 19.1% (14.7, 24.3) |
| Rwanda (2008) | 13.3% (10.8, 16.3) |  | 7.7% (5.8, 10.2%) | 11.0% (9.1, 13.2) |
| Seychelles (2015) | 24.1% (20.9, 27.5) |  | 13.6% (11.2, 16.3) | 18.9% (16.5, 21.5) |
| Somalia (2007) | 4.1% (2.6, 6.5) |  | 4.2% (2.4, 7.3) | 5.1% (3.3, 7.8) |
| Sudan (2009) | 15.4% (13.2, 17.8) |  | 6.7% (4.4, 10.0) | 11.9% (8.6, 16.3) |
| Tanzania (2016) | 6.2% (4.1, 9.3) |  | 3.1% (2.3, 4.2) | 5.2% (3.6, 7.3) |
| Uganda (2018) | 10.4% (8.1, 13.3) |  | 9.8% (7.3, 13.1) | 10.1% (8.4, 12.1) |
| **Regional Prevalence** | **10.9 (9.7, 12.2)** |  | **6.3% (5.5, 7.2)** | **8.9% (8.0, 9.9)** |
| **North Africa** |  |  |  |  |
| Algeria (2013) | 24.1% (21.1, 27.4) |  | 2.4% (1.7, 3.3) | 12.8% (11.3 to 14.5) |
| Egypt (2014) | 19.1% (13.7, 25.9) |  | 9.7% (5.2, 17.5) | 14% (10.5 to 19.9) |
| Libya (2010) | 13.2% (10.7, 16.1) |  | 5.8% (3.9, 8.6) | 9.8% (7.8 to 12.3) |
| Mauritania (2018) | 20.0% (16.3, 24.4) |  | 19.9% (13.8, 27.5) | 20.3% (16.0 to 25.4) |
| Morocco (2016) | 11.7% (9.1, 15.0) |  | 5.8% (4.1, 8.2) | 8.8% (7.0 to 11.0) |
| Tunisia (2017) | 18.7% (16.1, 21.5) |  | 5.1% (3.8, 6.8) | 11.9% (10.4 to13.6) |
| **Regional Prevalence** | **18.4% (15.3, 22.0)** |  | **7.3% (4.9, 10.9)** | **12.9% (10.7 to 15.5)** |
| **Southern Africa** |  |  |  |  |
| Angola (2010) | 1.7% (0.9, 3.1) |  | 0.4% (0.1, 1.4) | 1.2% (0.7, 2.0) |
| Botswana (2008) | 32.3% (28.3, 36.7) |  | 23.2% (20.0, 26.7) | 28.0% (25.1, 31.1) |
| Eswatini (2009) | 16.7% (13.6, 20.5) |  | 9.5% (7.4, 11.9) | 12.8% (10.7, 15.4) |
| Lesotho (2008) | 35.4% (29.4, 41.9) |  | 24.6% (21.2, 28.4) | 30.3% (26.5, 34.5) |
| Malawi (2009) | 15.6% (12.3, 19.5) |  | 11.8% (8.4, 16.4) | 14.5% (11.4, 18.2) |
| Mozambique (2013) | 7.6% (6.5, 9.0) |  | 8.1% (6.7, 9.7) | 8.2% (7.1, 9.5) |
| Namibia (2008) | 35.6% (31.1, 40.5) |  | 31.2% (27.1, 35.6) | 33.8% (30.1, 37.7) |
| South Africa (2011) | 40.4% (36.9, 44.1) |  | 30.0% (27.8, 32.3) | 35.2% (32.8, 37.8) |
| Zambia (2011) | 27.3% (24.2, 30.5) |  | 25.4% (23.0, 28.0) | 26.5% (23.5, 29.9) |
| Zimbabwe (2014) | 22.6% (17.4, 28.8) |  | 16.9% (12.9, 21.8) | 21.2% (16.9, 26.4) |
| **Regional Prevalence** | **29.7% (27.7, 32.0)** |  | **23.6% (22.1, 25.1)** | **27.1% (25.6, 28.6)** |
| **West Africa** |  |  |  |  |
| Benin (2003) | 14.4% (12.1,17.0) |  | 3.0% (2.0, 4.3) | 10.1% (8.7, 11.7) |
| Burkina Faso (2009) | 25.9% (22.7, 29.3) |  | 13.1% (11.0, 15.4) | 19.8% (17.5, 22.3) |
| Carbo Verde (2007) | 5.8% (3.9, 8.5) |  | 3.7% (2.5, 5.3) | 5.0% (3.7, 6.7%) |
| Cote D’Ivoire (2009) | 27.5% (24.0, 31.4) |  | 11.3% (8.9, 14.4) | 20.3% (17.4, 23.4) |
| Gambia (2017) | 18.1% (15.8, 20.7) |  | 5.8% (4.8, 7.0) | 11.8% (10.5, 13.3) |
| Ghana (2017) | 11.0% (8.5, 14.1) |  | 10.4% (7.0, 15.2) | 11.0% (8.30, 14.4) |
| Guinea (2008) | 34.3% (29.9, 39.0) |  | 21.3% (15.5, 28.4) | 29.9% (25.6, 34.5) |
| Guinea-Bissau (2008) | 10.8% (8.6, 13.5) |  | 9.8% (7.2, 13.3) | 10.3% (8.3, 12.8) |
| Liberia (2008) | 16.0% (11.3, 22.1) |  | 15.7% (11.7, 20.7) | 16.3% (12.1, 21.7) |
| Mali (2008) | 29.5% (23.7%, 36.1) |  | 15.1% (11.1, 20.3) | 23.4% (18.9, 28.7) |
| Niger (2008) | 8.9% (7.1, 11.1) |  | 0.8% (0.4, 1.7) | 4.9% (3.8, 6.2) |
| Nigeria (2009) | 19.2% (16.5, 22.2) |  | 16.9% (14.6, 19.5) | 18.9% (16.8, 21.2) |
| Senegal (2020) | 10.0% (8.4, 11.9) |  | 7.7% (5.8, 10.2) | 8.4% (7.2, 9.7) |
| Sierra Leone (2017) | 14.2% (10.9, 18.4) |  | 10.3% (7.8, 13.4) | 13.3% (10.0, 17.6) |
| Togo (2019) | 5.1% (4.1, 6.4) |  | 1.9% (1.2, 3.0) | 3.7% (3.0, 4.7) |
| **Regional prevalence** | **15.9% (14.6, 17.3)** |  | **10.4% (8.8, 12.1)** | **13.7% (12.5, 15.0)** |
| **Overall prevalence** | **17.4% (16.3, 18.6)** |  | **10.6% (9.5, 11.7)** | **14.3% (13.5, 15.3)** |

**Supplementary Table 4: Prevalence of use of different tobacco products by country and African region**

| Location | Cigarettes (95% CI) |  | Smokeless (95% CI) | Other smoked  (95% CI) | Shisha (95% CI) | Dual smokeless & smoked tobacco (95% CI) | Dual cigarettes & shisha (95% CI) |
| --- | --- | --- | --- | --- | --- | --- | --- |
| **Central Africa** |  |  |  |  |  |  |  |
| Burundi (2008) | 5.9% (3.7, 9.2) |  | 🞩 | 🞩 | 🞩 | 🞩 | 🞩 |
| Cameroon (2014) | 6.7% (4.2, 10.4) |  | 4.0% (2.9, 5.4) | 3.4% (2.3, 5.09) | 🞩 | 1.7% (1.0, 3.0) | 🞩 |
| CAR (2008) | 8.9% (7.1, 11.1) |  | 14.7% (12.4, 17.4) | 12.8% (5.9, 25.6) | 🞩 | 5.4% (4.4, 6.7) | 🞩 |
| Chad (2019) | 2.4% |  | 6.2% | 7.7% | 0.7% | 2.6% | 0.3% |
| Congo (2019) | 4.3% (3.2, 5.7) |  | 7.0% (5.9, 8.3) | 7.1% (5.4, 9.30) | 3.3% (2.3, 4.7) | 2.5% (2.0, 3.3) | 0.9% (0.6, 1.5) |
| DRC (2008) | 9.2% (7.2, 11.8) |  | 19.6% (17.2, 22.2) | 16.2% (12.9, 20.2) | 🞩 | 8.2% (6.8, 10.0) | 🞩 |
| Equatorial Guinea (2008) | 10.3% (8.1, 12.9) |  | 23.5% (18.0, 30.2) | 🞩 | 🞩 | 3.5% (2.2, 5.4) | 🞩 |
| Gabon (2014) | 9.1% (7.8, 10.6) |  | 3.2% (2.3, 4.3) | 3.9% (2.8, 5.5) | 2.7% (1.8, 4.0) | 1.0% (0.6, 1.6) | 1.3% (0.8, 2.1) |
| Sao Tome & Principe (2008) | 6.7% |  | 24.2% | 19.3% | 🞩 | 16.3% | 🞩 |
| **Regional Prevalence** | **6.9% (5.4, 8.7)** |  | **7.8% (6.8, 9.0)** | **6.7% (5.5, 8.2)** | **3.1% (2.3, 4.1)** | **3.2% (2.5, 3.9)** | **1.0% (0.8, 1.5)** |
| **East Africa** |  |  |  |  |  |  |  |
| Comoros (2015) | 11.9% (9.4, 15.0) |  | 8.9%(7.3, 10.7) | 🞩 | 🞩 | 4.2% (3.2, 5.6) | 🞩 |
| Djibouti (2013) | 7.7%(5.7, 10.4) |  | 7.8%(5.7, 10.0) | 6.1% (4.6, 8.2) | 12.5% (9.8, 15.8) | 4.5% (3.1, 6.4) | 2.9% (1.9, 4.4) |
| Eritrea (2006) | 2.0% (1.4, 2.7) |  | 🞩 | 🞩 | 🞩 | 🞩 | 🞩 |
| Ethiopia (2003) | 2.6% (1.7,3.9) |  | 🞩 | 🞩 | 🞩 | 🞩 | 🞩 |
| Kenya (2013) | 5.0% (3.8, 6.7) |  | 4.8% (3.9, 5.94) | 2.7% (2.0, 3.6) | 4.9% (3.7, 6.4) | 1.8% (1.2, 2.5) | 1.6% (1.1, 2.4) |
| Madagascar (2018) | 11.2% (9.1, 13.7) |  | 2.1% (1.3, 3.23) | 3.5% (2.2, 5.4) | 🞩 | 0.8% (0.4, 1.5) | 🞩 |
| Mauritius (2016) | 15.2% (10.9, 20.6) |  | 2.5% (2.1, 3.13) | 8.8% (6.3, 12.3) | 🞩 | 1.3% (0.8, 2.1) | 🞩 |
| Rwanda (2008) | 2.9% (2.1, 3.9) |  | 6.5% (5.2, 8%) | 6.0% (4.8, 7.5) | 🞩 | 3.6% (2.8, 4.6) | 🞩 |
| Seychelles (2015) | 15.6% (13.4, 18.2) |  | 1.9%(1.3, 2.6) | 9.8% (8.2, 11.7) | 13.6% (11.8, 15.7) | 1.3% (0.9, 2.0) | 5.9% (4.8, 7.2) |
| Somalia (2007) | 5.1% (3.3, 7.8) |  | 🞩 | 🞩 | 🞩 | 🞩 | 🞩 |
| Sudan (2009) | 3.6% (2.3, 5.4) |  | 5.6% (3.7, 8.3) | 🞩 | 7.4% (5.5, 10.0) | 🞩 | 1.8% (1.1, 3.0) |
| Tanzania (2016) | 1.4% (0.9, 2.1) |  | 2.6% (1.8, 3.77) | 2.9% (1.9, 4.5) | 🞩 | 2.6% (1.5, 4.3) | 🞩 |
| Uganda (2018) | 4.1% (2.6, 6.3) |  | 6.3% (5.2, 7.6) | 4.7% (3.2, 6.9) | 1.9% (1.1, 3.5) | 1.0% (0.6, 1.6) | 1.0% (0.5, 1.8) |
| **Regional Prevalence** | **4.2% (3.6, 4.8)** |  | **4.1% (3.6, 4.7)** | **3.4% (2.8, 4.2)** | **4.5% (3.7, 5.5)** | **1.5% (1.2, 1.8)** | **1.5% (1.1, 1.9)** |
| **North Africa** |  |  |  |  |  |  |  |
| Algeria (2013) | 9.2% (7.8, 10.7) |  | 5.6% (4.6, 6.8) | 5.6% (4.7, 6.5) | 🞩 | 3.9% (3.1, 4.8) | 🞩 |
| Egypt (2014) | 5.1% (2.9, 8.8) |  | 4.8% (2.9, 7.7) | 6.4% (3.7, 11.1) | 6.3% (4.3, 9.2) | 1.5% (1.0, 2.2) | 1.8% (0.9, 3.7) |
| Libya (2010) | 5.3% (3.8, 7.2) |  | 3.2% (2.1, 4.8) | 🞩 | 5.4% (4.1, 7.0) | 🞩 | 2.0% (1.3, 3.0) |
| Mauritania (2018) | 13.2% (9.0 to 19.0) |  | 6.9% (5.8, 8.3) | 7.9% (5.9, 10.6) | 🞩 | 3.0% (2.2, 4.1) | 🞩 |
| Morocco (2016) | 2.6% (1.9, 3.6) |  | 3.4% (2.5, 4.6) | 3.3% (2.4, 4.5) | 4.7% (3.1, 7.2) | 1.9% (1.2, 3.0) | 1.2% (0.7, 2.0) |
| Tunisia (2017) | 7.9% (6.4, 9.7) |  | 2.5% (1.8, 3.4) | 5.3% (4.6, 6.2) | 8.0% (6.7, 9.6) | 1.1% (0.7, 1.8) | 3.9% (3.0, 5.1) |
| **Regional Prevalence** | **5.6% (4.3, 7.3)** |  | **4.5% (3.4, 5.9)** | **5.6% (3.9, 7.9)** | **6.0% (4.6, 7.8)** | **2.0% (1.7, 2.5)** | **1.8% (1.2, 2.9)** |
| **Southern Africa** |  |  |  |  |  |  |  |
| Angola (2010) | 1.2% (0.7, 2.0) |  | 🞩 | 🞩 | 🞩 | 🞩 | 🞩 |
| Botswana (2008) | 17.8% (15.0, 21.0) |  | 12.8%(11.0, 14.8) | 9.4% (8.1, 10.9) | 🞩 | 7.1% (5.9, 8.7) | 🞩 |
| Eswatini (2009) | 6.6% (4.7, 9.3) |  | 6.7% (5.0, 9.0) | 4.9% (4.0, 5.93) | 🞩 | 3.7% (2.9, 4.6) | 🞩 |
| Lesotho (2008) | 12.8% (9.6, 16.8) |  | 17.1% (14.7, 19.9) | 14.5% (12.4, 16.0) | 🞩 | 9.0% (7.4, 11.0) | 🞩 |
| Malawi (2009) | 3.3% (2.1, 5.2) |  | 9.3% (6.6, 12.9) | 8.4% (6.5, 10.8) | 🞩 | 4.5% (2.9, 7.1) | 🞩 |
| Mozambique (2013) | 2.4% (1.9, 3.07%) |  | 4.3% (3.6, 5.1) | 4.2% (3.4, 5.2) | 🞩 | 1.5% (1.1, 2.0) | 🞩 |
| Namibia (2008) | 12.3% (9.7, 15.5) |  | 19.4% (16.1, 23.3) | 17.9% (14.9, 21.5) | 🞩 | 9.4% (6.9, 12.9) | 🞩 |
| South Africa (2011) | 16.4% (14.5,18.6) |  | 16.0%(14.8, 17.3) | 18.6% (16.9, 20.3) | 🞩 | 9.7% (8.6, 10.9) | 🞩 |
| Zambia (2011) | 7.0% (5.5, 9.0) |  | 15.1% (12.6, 18) | 14.6% (12.8, 16.5) | 🞩 | 5.5% (4.2, 7.1) | 🞩 |
| Zimbabwe (2014) | 17.8% (13.4, 23.4) |  | 7.3% (4.9, 10.7) | 6.7% (4.9, 9.3) | 🞩 | 3.1% (1.8, 5.1) | 🞩 |
| **Regional Prevalence** | **12.7% (11.4, 14.0)** |  | **12.8% (12.0, 13.7)** | **14.0% (13.0, 15.0)** | 🞩 | **7.3% (6.6, 8.0)** | 🞩 |
| **West Africa** |  |  |  |  |  |  |  |
| Benin (2003) | 10.1% (8.7, 11.7) |  | 🞩 | 🞩 | 🞩 | 🞩 | 🞩 |
| Burkina Faso (2009) | 7.6% (5.9, 9.7) |  | 11.3% (10.0, 12.8) | 7.1% (6.2, 8.2) | 🞩 | 3.9% (3.3, 4.7) | 🞩 |
| Carbo Verde (2007) | 5.0% (3.7, 6.7) |  | 🞩 | 🞩 | 🞩 | 🞩 | 🞩 |
| Cote D’Ivoire (2009) | 15.2% (12.8, 18.1) |  | 5.5% (4.4, 7.0) | 4.5% (3.6, 5.6) | 🞩 | 2.1% (1.6, 2.8) | 🞩 |
| Gambia (2017) | 8.5% (7.3, 9.8) |  | 1.8% (1.5, 2.2) | 4.7% (3.7, 5.9) | 🞩 | 0.9% (0.7, 1.2) | 🞩 |
| Ghana (2017) | 3.0% (2.1, 4.3) |  | 3.6% (2.6, 4.9) | 4.4% (3.1, 6.1) | 6.2% (4.3, 8.9) | 1.7% (1.1, 2.9) | 1.0% (0.5,1.9) |
| Guinea (2008) | 11.4% (8.4, 15.4) |  | 19.2% (15.7, 24.1) | 11.8% (10.2, 13.6) | 🞩 | 7.8% (6.3, 9.7) | 🞩 |
| Guinea-Bissau (2008) | 4.3% (3.4, 5.4) |  | 4.1% (2.7, 6.2) | 4.8% (3.4, 6.9) | 🞩 | 2.5% (1.4, 4.4) | 🞩 |
| Liberia (2008) | 2.2% (1.3, 3.8) |  | 10.8% (7.7, 14.9) | 11.2% (8.0, 15.5) | 🞩 | 6.1% (3.9, 9.3) | 🞩 |
| Mali (2008) | 11.7% (8.8%, 15.4) |  | 8.4% (5.5, 12.5) | 10.2% (7.7,o 13.6) | 🞩 | 3.3% (2.1, 5.0) | 🞩 |
| Niger (2008) | 4.9% (3.8, 6.2) |  | 🞩 | 🞩 | 🞩 | 🞩 | 🞩 |
| Nigeria (2009) | 4.1% (3.1, 5.6) |  | 11.2% (9.8, 12.8) | 10.9% (9.4, 12.4) | 🞩 | 5.2% (4.4, 6.2) | 🞩 |
| Senegal (2020) | 3.9% (3.3, 4.7) |  | 3.3% (2.4, 4.7) | 5.2(4.4, 6.2) | 2.6% (2.1, 3.2) | 1.3% (0.9, 1.8) | 1.0% (0.7, 1.5) |
| Sierra Leone (2017) | 4.0% (2.9, 5.7) |  | 6.8% (3.7, 12.21) | 5.9% (3.6, 9.4) | 6.4% (4.6, 8.8) | 4.3% (2.0, 9.1) | 1.3% (0.8, 2.0) |
| Togo (2019) | 3.2% (2.4, 4.3) |  | 1.54% (1.1, 2.08) | 2.2% (1.6, 2.9) | 1.5% (1.0, 2.2) | 0.7% (0.4, 1.0) | 0.8% (0.5, 1.3) |
| **Regional Prevalence** | **5.5% (4.9, 6.1)** |  | **6.3% (5.5, 7.16)** | **6.2% (5.5, 6.9)** | **4.7% (3.6, 6.1)** | **2.8% (2.4, 3.3)** | **1.0% (0.7, 1.4)** |
| **Overall Prevalence** | **6.4% (5.9, 7.0)** |  | **6.4% (5.9, 6.9)** | **6.7% (6.0, 7.4)** | **5.2% (4.4, 6.1)** | **3.0% (2.8, 3.2)** | **1.5% (1.2, 2.0)** |

*Notes:* The estimates for Chad and Sao Tome & Principe are not represented with 95 CIs because they represent population estimates. 🞩 indicates that data is not available.

**Supplementary Table 5: Prevalence of different of tobacco products among boys, by country and African region**

|  | **Cigarettes (95% CI)** | **Other smoked tobacco (95% CI)** | **Smokeless tobacco (95% CI)** | | **Shisha(95% CI)** | **Dual use of shisha & cigarettes(95% CI)** |  | **Dual use of smoked and smokeless (95% CI)** |
| --- | --- | --- | --- | --- | --- | --- | --- | --- |
| **Central Africa** | **Panel A: Boys** | | | | | | |  |
| Burundi (2008) | 8.5% (4.9, 14.5) | 🞩 | 🞩 | 🞩 | | 🞩 |  | 🞩 |
| Cameroon (2014) | 9.6% (6.1, 14.9) | 4.3% (2.9, 6.1) | 5.2% (3.7, 7.1) | 🞩 | | 🞩 |  | 2.3% (1.3, 4.1) |
| CAR (2008) | 11.9% (9.0, 15.5) | 9.4% (7.4, 12.0) | 18.6% (14.6, 23.4) | 🞩 | | 🞩 |  | 6.9% (5.3, 8.8) |
| Chad (2019) | 27.4% | 7.0% | 5.4% | 0.5% | | 0.2% |  | 2.4% |
| Congo (2019) | 5.6% (4.0, 7.7) | 7.9% (5.6, 10.9) | 7.9% | 2.4% (1.8, 3.3) | | 1.1% (0.6, 1.9) |  | 3.1% (2.3, 4.2) |
| DRC (2008) | 12.2% (9.5, 15.5) | 17.7% (13.4, 23.2) | 20.7% (17.4, 24.6) | 🞩 | | 🞩 |  | 8.5% (6.2, 11.5) |
| Equatorial Guinea (2008) | 13.8% (10.5, 18.1) | 🞩 | 25.9% (19.5, 33.6) | 🞩 | | 🞩 |  | 5.1% (3.2, 7.9) |
| Gabon (2014) | 13.2% (11.7, 14.8) | 4.4% (2.8, 6.8) | 3.6% (2.0, 6.1) | 2.7% (1.5, 4.8) | | 1.5% (0.8, 2.8) |  | 1.2% (0.6, 2.4) |
| **Regional prevalence** | **9.6% (7.5, 12.3)** | **7.4% (6.1, 8.9%)** | **8.8% (7.6, 10.1)** | **2.5% (1.9, 3.3)** | | **1.2% (0.8, 1.8)** |  | **3.6% (2.9, 4.6)** |
| **East Africa** |  |  |  |  | |  |  |  |
| Comoros (2015) | 14.7% (11.4, 18.8) | 🞩 | 11.5% (9.5, 13.9) | 🞩 | | 🞩 |  | 6.1% (4.6, 8.0) |
| Djibouti (2013) | 10.5% (7.6, 14.3) | 6.4% (4.3, 9.3) | 9.7% (6.9, 13.3) | 11.1% (8.5, 14.3) | | 3.6% (2.5, 5.3) |  | 5.8% (3.8, 8.8) |
| Eritrea (2006) | 2.6% (2.0, 3.5) | 🞩 | 🞩 | 🞩 | | 🞩 |  | 🞩 |
| Ethiopia (2003) | 3.5% (2.4, 5.2) | 🞩 | 🞩 | 🞩 | | 🞩 |  | 🞩 |
| Kenya (2013) | 6.8% (4.7, 9.8) | 3.0% (2.0, 4.4) | 5.8% (4.4, 7.5) | 4.6% (3.2, 6.6) | | 1.5% (0.9, 2.6) |  | 2.1% (1.2, 3.5) |
| Madagascar (2018) | 18.7% (14.9, 23.2) | 5.3% (3.5, 7.9) | 2.8% (1.6, 4.7) | 🞩 | | 🞩 |  | 1.6% (0.8, 2.9) |
| Mauritius (2016) | 23.4% (17.6, 30.6) | 13.5% (10.3, 17.5) | 2.6% (1.9, 3.7) | 🞩 | | 🞩 |  | 1.5% (0.9, 2.5) |
| Rwanda (2008) | 4.3% (3.1, 5.9) | 7.2% (5.4, 9.4) | 6.9% (5.3, 8.9) | 🞩 | | 🞩 |  | 3.9% (2.9, 5.2) |
| Seychelles (2015) | 20.3% (17.2, 23.9) | 12.3% (10.0, 14.9) | 2.9% (2.0, 4.4) | 17.2% (14.8, 20.0) | | 8.6% (6.8, 10.8) |  | 2.2% (1.4, 3.4) |
| Somalia (2007) | 4.1% (2.6, 6.5) | 🞩 | 🞩 | 🞩 | | 🞩 |  | 🞩 |
| Sudan (2009) | 4.8% (3.4, 6.7) | 🞩 | 7.4% (5.6, 9.5) | 9.5% (7.8, 11.5) | | 2.6% (1.7, 3.8) |  | 3.4% (2.1, 5.3) |
| Tanzania (2016) | 1.8% (1.1, 3.0) | 3.7% (2.2, 6.0) | 3.1% (2.0, 4.7) | 🞩 | | 🞩 |  | 1.5% (0.8, 2.6) |
| Uganda (2018) | 5.0% (3.1, 7.9) | 4.7% (2.9, 7.7) | 7.0% (5.4, 9.0) | 2.6% (1.4, 4.9) | | 1.6% (0.7, 3.3) |  | 2.2% (1.3, 3.7) |
| **Regional prevalence** | **6.0% (5.1, 7.0)** | **4.1% (3.2, 5.2)** | **5.0% (4.3, 5.7)** | **5.0% (4.0, 6.3)** | | **1.8% (1.3, 2.5)** |  | **2.0% (1.6, 2.6)** |
| **North Africa** |  |  |  |  | |  |  |  |
| Algeria (2013) | 18.2% (15.5, 21.2) | 10.2% (8.7, 11.9) | 11.2% (9.2, 13.7) | 🞩 | | 🞩 |  | 7.9% (6.4, 9.8) |
| Egypt (2014) | 8.8% (4.3, 17.3) | 10.3% (5.4, 18.6) | 4.9% (3.3, 7.2) | 8.6% (5.9, 12.4) | | 2.8% (1.1, 7.2) |  | 2.2% (1.2, 4.1) |
| Libya (2010) | 7.7% (5.6, 10.4) | 🞩 | 3.5% (2.3, 5.1) | 7.6% (5.7, 9.9) | | 3.2% (2.1, 4.7) |  | 2.1% (1.4, 3.1) |
| Mauritania (2018) | 13.3% (9.8, 17.7) | 7.6% (5.4, 10.5) | 6.6% (5.2, 8.4) | 🞩 | | 🞩 |  | 2.9% (2.1, 4.0) |
| Morocco (2016) | 4.0% (2.5, 6.4) | 3.9% (2.6, 5.6) | 4.8% (3.2, 7.2) | 7.1% (4.7, 10.6) | | 2.2% (1.2, 3.9) |  | 2.4% (1.4, 4.1) |
| Tunisia (2017) | 14.6% (11.8, 17.9) | 9.0% (7.4, 10.8) | 3.4% (2.4, 4.7) | 13.6% (11.3, 16.2) | | 7.7% (5.8, 10.1) |  | 1.8% (1.1, 2.8) |
| **Regional prevalence** | **10.0% (7.1, 13.8)** | **8.9% (5.9, 13.2)** | **5.9% (4.9, 7.1)** | **8.6% (3.8, 11.1)** | | **3.1% (1.7, 5.4)** |  | **3.2% (2.5, 4.2)** |
| **Southern Africa** |  |  |  |  | |  |  |  |
| Angola (2010) | 1.7% (0.9, 3.1) | 🞩 | 🞩 | 🞩 | | 🞩 |  | 🞩 |
| Botswana (2008) | 21.8% (17.9, 26.3) | 11.9% (10.0, 14.1) | 12.4% (10.0, 15.3) | 🞩 | | 🞩 |  | 7.1% (5.4, 9.2) |
| Eswatini (2009) | 9.2% (6.7, 12.50 | 6.5% (4.5, 9.3) | 8.1% (5.2, 12.5) | 🞩 | | 🞩 |  | 4.7% (3.0, 7.3) |
| Lesotho (2008) | 17.8% (12.9, 24.0) | 16.7% (14.6, 18.9) | 18.9% (14.9, 23.7) | 🞩 | | 🞩 |  | 10.7% (8.8, 13.1) |
| Malawi (2009) | 4.2% (2.2, 7.6) | 8.4% (6.2, 11.3) | 9.9% (14.9, 23.7) | 🞩 | | 🞩 |  | 4.9% (2.9, 8.3) |
| Mozambique (2013) | 1.9% (1.4, 2.6) | 3.8% (3.0, 4.8) | 4.5% (3.6, 5.5) | 🞩 | | 🞩 |  | 1.5% (1.0, 2.1) |
| Namibia (2008) | 12.7% (9.1, 17.6) | 19.0% (14.8, 24.2) | 20.6% (16.2, 25.8) | 🞩 | | 🞩 |  | 10.3% (6.4, 16.1) |
| South Africa (2011) | 21.5% (18.8, 24.4) | 21.0% (18.5, 23.8) | 17.6% (15.6, 19.8) | 🞩 | | 🞩 |  | 11.7% (9.9, 13.8) |
| Zambia (2011) | 7.8% (6.5, 9.3) | 14.8% (13.4, 16.3) | 15.6% (12.0, 20.1) | 🞩 | | 🞩 |  | 5.9% (3.8, 9.0) |
| Zimbabwe (2014) | 17.8% (12.7, 24.5) | 9.0% (5.5, 14.3) | 7.4% (4.4, 12.2) | 🞩 | | 🞩 |  | 2.7% (1.8, 3.9) |
| **Regional prevalence** | **15.4% (13.7, 17.2)** | **15.3% (13.8, 17.0)** | **13.7% (12.4, 15.2)** | 🞩 | | 🞩 |  | **8.3% (7.3, 9.6)** |
| **West Africa** |  |  |  |  | |  |  |  |
| Benin (2003) | 14.4% (12.1, 17.0) | 🞩 | 🞩 | 🞩 | | 🞩 |  | 🞩 |
| Burkina Faso (2009) | 12.7% (10.1, 15.8) | 9.4% (8.0, 11.0) | 13.2% (11.5, 15.0) | 🞩 | | 🞩 |  | 5.1% (4.0, 6.4) |
| Carbo Verde (2007) | 5.8% (3.9, 8.5) | 🞩 | 🞩 | 🞩 | | 🞩 |  | 🞩 |
| Cote D’Ivoire (2009) | 23.4% (20.0, 27.2) | 5.7% (4.4, 7.5) | 5.5% (4.2, 7.20 | 🞩 | | 1.4% (0.9, 2.2) |  | 2.4% (1.8, 3.2) |
| Gambia (2017) | 14.1% (11.9, 16.6) | 7.2% (5.5, 9.3) | 2.6% (2.0, 3.3) | 🞩 | | 🞩 |  | 1.5% (1.0, 2.1) |
| Ghana (2017) | 3.2% (2.5, 4.2) | 4.5% (3.2, 6.4) | 3.2% (2.3, 4.4) | 5.8% (4.1, 8.2) | | 0.6% (0.4, 1.1) |  | 1.4% (0.9, 2.1) |
| Guinea (2008) | 15.7% (12.1, 20.2) | 13.2% (11.2, 15.6) | 20.8% (16.5, 25.8) | 🞩 | | 🞩 |  | 9.1% (7.0, 11.9) |
| Guinea-Bissau (2008) | 5.9% (4.4, 7.8) | 3.4% (2.2, 5.4) | 3.5% (2.1, 5.9) | 🞩 | | 🞩 |  | 1.9% (1.0, 3.3) |
| Liberia (2008) | 2.6% (1.4, 4.7) | 11.7% (7.9, 17.2) | 9.9% (6.6, 14.7) | 🞩 | | 🞩 |  | 6.0% (3.6, 9.8) |
| Mali (2008) | 18.4% (13.9, 24.0) | 11.6% (8.2, 16.3) | 9.6% (6.5,14.0) | 🞩 | | 🞩 |  | 4.3% (2.6, 7.0) |
| Niger (2008) | 8.9% (7.1, 11.2) | 🞩 | 🞩 | 🞩 | | 🞩 |  | 🞩 |
| Nigeria (2009) | 5.0% (3.5, 7.0) | 10.8% (8.7, 13.3) | 11.1% (9.4, 13.0) | 🞩 | | 🞩 |  | 4.7% (3.7, 6.0) |
| Senegal (2020) | 6.3% (4.9, 8.0) | 6.3% (5.0, 7.8) | 3.9% (2.7, 5.6) | 3.2% (2.4, 4.3) | | 1.6% (1.1, 2.3) |  | 1.0% (0.7, 1.4) |
| Sierra Leone (2017) | 6.5% (4.6, 9.0) | 7.2% (4.7, 10.9) | 6.6% (3.9, 10.9) | 7.5% (5.3, 10.6) | | 2.2% (1.4, 3.6) |  | 1.8% (0.6, 5.1) |
| Togo (2019) | 5.3% (3.9, 7.1) | 3.2% (2.3, 4.4) | 2.0% (1.5, 2.6) | 2.4% (1.6, 3.5) | | 1.4% (0.9, 2.2) |  | 0.2 (0.1, 0.5) |
| **Regional prevalence** | **8.0% (7.2, 8.8)** | **7.0% (6.3, 7.8)** | **6.7% (5.9, 7.7)** | **4.8% (3.6, 6.1)** | | **1.1% (0.9, 1.4)** |  | **3.1% (2.6, 3.7)** |
| **Overall (Africa) Prevalence** | **9.2% (8.2, 10.4)** | **8.3% (7.2, 9.5)** | **7.3% (6.8, 7.8)** | **3.4% (3.0, 3.8)** | | **2.2% (1.6, 3.1)** |  | **3.8% ( 3.4, 4.1)** |

*Notes:* 🞩 indicates that data is not available

**Supplementary Table 6: Prevalence of different of tobacco products among girls, by country and African region**

|  | **Cigarettes (95% CI)** | **Other smoked tobacco (95% CI)** | **Smokeless tobacco (95% CI)** | **Shisha(95% CI)** | **Dual use of shisha & cigarettes(95% CI)** | **Dual use of smoked and smokeless (95% CI)** |
| --- | --- | --- | --- | --- | --- | --- |
| **Central Africa** |  |  |  |  |  |  |
| Burundi (2008) | 2.7% (1.5, 5.0) | 🞩 | 🞩 | 🞩 | 🞩 | 🞩 |
| Cameroon (2014) | 3.2% (2.0, 5.0) | 2.4% (1.3, 4.5) | 2.6% (1.8, 3.7) | 🞩 | 🞩 | 1.0% (0.5, 2.1) |
| CAR (2008) | 4.4% (3.0, 6.40 | 16.0% (4.8, 4.2) | 8.8% (6.6, 1.2) | 🞩 | 🞩 | 2.5% (1.8, 3.4) |
| Chad (2019) | 1.6% | 8.0% | 6.3% | 0.7%(0.7 to 0.7) | 0.2% (0.15, 0.15) | 2.4% |
| Congo (2019) | 2.5% (1.7, 3.5) | 5.6% (4.1, 7.5) | 5.6% (4.5, 6.9) | 2.9%(1.9 to 4.4) | 0.6%(0.3 to 1.1) | 1.6% (1.1, 2.5) |
| DRC (2008) | 5.0% (3.6, 6.9) | 14.0% (10.2, 18.6) | 17.7% (14.3, 21.7) | 🞩 | 🞩 | 7.0% (5.2, 9.4) |
| Equatorial Guinea (2008) | 6.1% (4.6, 8.1) | 🞩 | 20.2% (14.6, 27.2) | 🞩 | 🞩 | 1.7% (1.0, 3.1) |
| Gabon (2014) | 4.7% (3.2, 6.9) | 3.4% (2.2, 5.2) | 2.7% (2.0, 3.7) | 2.6%(1.5 to 4.5) | 1.1%(0.6 to 2.0) | 0.8% (0.5, 1.3) |
| **Regional prevalence** | **3.4% (2.7, 4.4)** | **5.3% (4.0, 7.0)** | **6.1% (5.0, 7.4)** | **2.8%(2.0 to 3.9)** | **0.8%(0.5 to 1.2)** | **2.2% (1.6, 3.0)** |
| **East Africa** |  |  |  |  |  |  |
| Comoros (2015) | 9.1% (7.0, 11.8) | 🞩 | 6.1% (4.7, 7.9) | 🞩 | 🞩 | 2.4% (1.6, 3.5) |
| Djibouti (2013) | 3.6% (1.8, 7.2) | 5.5% (3.5, 8.5) | 4.9% (3.5, 6.8) | 14.1% (10.6, 18.8) | 1.9%(0.7 to 4.8) | 2.8% (1.7, 4.5) |
| Eritrea (2006) | 0.7% (3.4, 1.5) | 🞩 | 🞩 | 🞩 | 🞩 | 🞩 |
| Ethiopia (2003) | 1.1% (0.5, 2.3) | 🞩 | 🞩 | 🞩 | 🞩 | 🞩 |
| Kenya (2013) | 3.1% (2.1, 4.5) | 2.0% (1.2, 3.2) | 3.7% (2.9, 4.8) | 5.0% (3.8, 6.6) | 1.8%(1.1 to 3.1) | 1.3% (0.8, 2.2) |
| Madagascar (2018) | 4.1% (3.0, 5.7) | 1.6% (0.7, 3.6) | 1.4% (0.6, 3.2) | 🞩 | 🞩 | 0.1% (0.0, 0.5) |
| Mauritius (2016) | 7.4% (4.6, 11.8) | 4.4% (2.5, 7.5) | 2.5% (1.7, 3.6) | 🞩 | 🞩 | 1.1% (0.5, 2.5) |
| Rwanda (2008) | 1.0% (0.5, 2.1) | 4.3% (3.1, 6.1) | 5.5% (4.1, 7.4) | 🞩 | 🞩 | 2.9% (2.0, 4.3) |
| Seychelles (2015) | 11.1% (9.0, 13.6) | 7.4% (5.7, 9.5) | 0.8% (0.4, 1.4) | 9.8% (7.9, 12.2) | 3.2%(2.4 to 4.4) | 0.4% (0.2, 1.0) |
| Somalia (2007) | 4.2% (2.4, 7.3) | 🞩 | 🞩 | 🞩 | 🞩 | 🞩 |
| Sudan (2009) | 2.0% (2.4, 7.3) | 🞩 | 2.2% (1.0, 4.5) | 4.4% (3.1, 6.4) | 0.9%(0.4 to 1.9) | 1.0% (0.5, 2.0) |
| Tanzania (2016) | 0.7% (0.3, 1.3) | 1.5 (0.9, 2.6) | 1.8% (1.1, 2.8) | 🞩 | 🞩 | 0.4% (0.2, 1.0) |
| **Regional prevalence** | **2.2% (1.8, 2.7)** | **2.4% (1.8, 3.1)** | **3.0% (2.4, 3.6)** | **3.6% (2.9, 4.4)** | **1.1%(0.7 to 1.7)** | **0.7% (0.6, 1.1)** |
| **North Africa** |  |  |  |  |  |  |
| Algeria (2013) | 1.1% (0.7, 1.7) | 1.4% (1.0, 1.8) | 0.4% (0.2, 0.7) | 🞩 | 🞩 | 0.1% (0.0, 0.3) |
| Egypt (2014) | 1.4% (0.7, 3.0) | 2.5% (1.2, 5.2) | 4.5% (1.7, 11.4) | 3.8%(1.9 to 7.3) | 0.8%(0.3 to 2.0) | 0.5% (0.1, 1.9) |
| Libya (2010) | 2.4% (1.2, 4.7) | 🞩 | 2.5% (1.5, 4.1) | 3.2%(2.1 to 4.8) | 0.8%(0.4 to 1.9) | 1.5% (0.7, 3.0) |
| Mauritania (2018) | 12.5% (7.0, 21.3) | 8.0% (5.6, 11.3) | 7.0% (5.2, 9.5) | 🞩 | 🞩 | 3.1% (1.9, 4.9) |
| Morocco (2016) | 1.0% (0.7, 1.6) | 2.7% (1.8, 4.0) | 2.1% (1.3, 3.4) | 2.6%(1.4 to 4.9) | 0.2% (0.05, 0.64) | 1.5% (0.9, 2.4) |
| Tunisia (2017) | 1.3% (0.8, 2.2) | 1.6% (0.9, 2.7) | 1.6% (1.0, 2.6) | 2.4%(1.6 to 3.7) | 0.16%(0.0 to 0.6) | 0.5% (0.2, 1.3) |
| **Regional prevalence** | **1.5% (1.0, 2.2)** | **2.3 (1.5, 3.6)** | **3.0% (1.5, 6.2)** | **3.4%(2.1 to 5.4)** | **0.6%(0.3 to 1.3)** | **0.7% (0.4, 1.2)** |
| **Southern Africa** |  |  |  |  |  |  |
| Angola (2010) | 0.4% (0.1, 1.40) | 🞩 | 🞩 | 🞩 | 🞩 | 🞩 |
| Botswana (2008) | 13.3% (10.3, 17.0) | 7.1% (5.8, 8.7) | 12.7% (10.6, 15.1) | 🞩 | 🞩 | 6.9% (5.4, 8.8) |
| Eswatini (2009) | 4.5% (2.9, 7.0) | 3.6% (2.4, 5.2) | 5.4% (4.4, 6.6) | 🞩 | 🞩 | 2.8% (2.0, 4.1) |
| Lesotho (2008) | 8.3% (6.1, 11.1) | 11.9% (9.8, 14.5) | 15.3% ( 11.9, 19.5) | 🞩 | 🞩 | 7.6% (5.8, 9.9) |
| Malawi (2009) | 1.6% (0.9, 2.8) | 7.5% (5.3, 10.5) | 7.9% (5.3, 11.4) | 🞩 | 🞩 | 3.9% (2.6, 6.0) |
| Mozambique (2013) | 2.7% (2.0. 3.7) | 4.2% (3.3, 5.4) | 3.9% (3.0, 4.9) | 🞩 | 🞩 | 1.5% (1.0, 2.2) |
| Namibia (2008) | 11.2% (8.4, 14.8) | 16.6% (14.0, 19.5) | 17.6% (14.2, 21.4) | 🞩 | 🞩 | 8.1% (6.2, 10.6) |
| South Africa (2011) | 11.9% (9.9, 14.1) | 16.2% (14.6, 17.8) | 13.8% (12.4, 15.4) | 🞩 | 🞩 | 7.7% (6.5, 9.0) |
| Zambia (2011) | 5.8% (5.6, 7.3) | 14.0% (12.4, 15.6) | 14.4% (13.4, 15.4) | 🞩 | 🞩 | 5.0% (4.9, 5.2) |
| Zimbabwe (2014) | 13.4% (9.6, 18.5) | 4.3% (3.2, 5.8) | 7.5% (4.4, 12.5) | 🞩 | 🞩 | 3.5% (1.4, 8.6) |
| **Regional prevalence** | **9.6% (8.3, 11.0)** | **12.4 (11.4, 13.4)** | **11.5% (10.5, 12.6)** | 🞩 | 🞩 | **6.1% (5.3, 7.0)** |
| **West Africa** |  |  |  |  |  |  |
| Benin (2003) | 3.0% (2.0, 4.3) | 🞩 | 🞩 | 🞩 | 🞩 | 🞩 |
| Burkina Faso (2009) | 2.3% (1.2, 4.3) | 4.7% (3.7, 5.8) | 9.6% (8.0, 11.3) | 🞩 | 🞩 | 3.0% (2.3, 4.0) |
| Carbo Verde (2007) | 3.7% (2.5, 5.3) | 🞩 | 🞩 | 🞩 | 🞩 | 🞩 |
| Cote D’Ivoire (2009) | 5.6% (4.0, 7.9) | 2.9% (2.0, 4.2) | 5.7% (4.2, 7.7) | 🞩 | 🞩 | 1.9% (1.2, 2.9) |
| Gambia (2017) | 3.3% (2.5, 4.3) | 2.3% (1.7, 3.1) | 1.1% (0.8, 1.4) | 🞩 | 🞩 | 0.3% (0.2, 0.4) |
| Ghana (2017) | 2.7% (1.3, 5.3) | 3.7% (2.2, 6.1) | 4.0% (2.5, 6.2) | 6.2%(3.7 to 10.1) | 1.4%(0.6 to 3.2) | 2.1% (1.0, 4.3) |
| Guinea (2008) | 4.0% (2.3, 6.9) | 8.2% (6.1, 11.0) | 15.6% (10.3, 22.8) | 🞩 | 🞩 | 4.9% (3.1, 7.7) |
| Guinea-Bissau (2008) | 2.6% (1.5, 4.5) | 6.3% (3.8, 10.4) | 4.7% (2.8, 7.8) | 🞩 | 🞩 | 3.2% (1.6, 6.2) |
| Liberia (2008) | 1.2% (0.5, 2.6) | 10.2% (7.4, 13.8) | 10.8% (7.4, 15.6) | 🞩 | 🞩 | 5.6% (3.4, 9.2) |
| Mali (2008) | 3.2% (2.0, 5.1) | 8.5% (6.3, 11.3) | 6.2% (3.4, 11.0) | 🞩 | 🞩 | 1.9% (1.1, 3.2) |
| Niger (2008) | 0.8% (0.4, 1.7) | 🞩 | 🞩 | 🞩 | 🞩 | 🞩 |
| Nigeria (2009) | 2.9% (2.2, 4.0) | 9.9% (8.4, 11.8) | 10.4% (8.5, 12.6) | 🞩 | 🞩 | 4.7% (3.7, 6.0) |
| Senegal (2020) | 1.7% (1.1, 2.5) | 4.3% (3.2, 5.7) | 2.7% (1.9, 3.8) | 1.9%(1.4 to 2.5) | 0.5%(0.2 to 1.1) | 1.0% (0.7, 1.4) |
| Sierra Leone (2017) | 1.5% (0.8, 2.6) | 2.9% (1.9, 4.4) | 5.1% (2.8, 9.0) | 5.0%(3.2 to 7.5) | 0.3%(0.1 to 0.7) | 1.8% (0.6, 5.1) |
| Togo (2019) | 6.7% (0.3, 1.3) | 0.9% (0.5, 1.6) | 1.0% (0.5, 1.8) | 0.4%(0.2 to 1.0) | 0.1%(0.0 to 0.5) | 0.2% (0.1, 0.5) |
| **Regional prevalence** | **2.5% (1.9, 3.3)** | **4.6% (3.9, 5.5)** | **5.3% (4.4, 6.3)** | **4.2%(2.8 to 6.2)** | **0.9%(0.4 to 1.7)** | **2.1% (1.6, 2.8)** |
| **Overall prevalence** | **3.4%** | **4.8% (4.3, 5.4)** | **5.1% (4.4, 5.9)** | **3.6%(2.8 to 4.6)** | **0.8%(0.6 to 1.1)** | **2.0% (1.8, 2.3)** |

*Notes:* 🞩 indicates that data is not available

**Supplementary Table 7: Robustness checks of the prevalence of different tobacco products by gender, age group, African region, and World Bank income group (2010 – 2020)**

|  | **Cigarettes (95% CI)** | **Other smoked tobacco (95% CI)** | **Smokeless tobacco (95% CI)** | **Shisha (95% CI)** | **Dual use of shisha & cigarettes (95% CI)** | **Dual use of smoked & smokeless (95% CI)** |
| --- | --- | --- | --- | --- | --- | --- |
| **African region** |  |  |  |  |  |  |
| Central Africa | 6.4% (4.5, 8.7) | 4.3% (3.3, 5.6) | 4.6% (3.9, 5.6) | 3.1% (2.3, 4.1) | 1.0% (0.7, 1.5) | 1.9% (1.3, 2.8) |
| East Africa | 4.4% (3.8, 5.2) | 3.3% (2.7, 4.1) | 3.9% (3.4, 4.5) | 3.6% (2.8, 4.7) | 1.3% (1.0, 1.9) | 1.3% (1.0, 1.6) |
| North Africa | 5.6% (5.3, 7.3) | 5.6% (3.9, 7.9) | 4.5% (3.4, 5.9) | 6.0% (4.6, 7.8) | 1.8% (1.2, 2.9) | 2.0% (1.7, 2.5) |
| Southern Africa | 14.1% (12.6, 15.7) | 14.9% (13.8, 16.2) | 13.1% (12.2, 14.1) | 🞩 | 🞩 | 7.7% (6.9, 8.5) |
| West Africa | 3.5% (2.9, 4.2) | 4.4% (3.6, 5.3) | 3.4% (2.8, 4.2) | 4.7% (3.6, 6.1) | 1.0% (0.7, 1.4) | 1.7% (1.2, 2.3) |
| **World Bank Income Group** |  |  |  |  |  |  |
| High income | 15.6% (13.4, 18.2) | 9.8% (8.2, 11.7) | 1.9% (1.3, 2.6) | 13.6% (11.8, 15.6) | 5.9% (4.8, 7.2) | 1.3% (0.9, 2.0) |
| Upper middle income | 15.0% (13.3, 16.8) | 17.9% (16.3, 19.5) | 14.1% (13.1, 15.1) | 4.7% (3.7, 5.9) | 1.8% (1.3, 2.6) | 8.5% (7.6, 9.5) |
| Lower middle income | 4.8% (4.1, 5.7) | 4.6% (3.7, 5.7) | 4.2% (3.6, 4.9) | 5.6% (4.6, 6.8) | 1.6% (1.1, 2.2) | 1.8% (1.6, 2.1) |
| Low income | 5.7% (4.8, 6.6) | 4.1% (3.3, 5.0) | 4.3% (3.7, 4.9) | 4.7% (3.7, 5.9) | 1.0% (0.6, 1.5) | 1.4% (1.1, 1.8) |
| **Age Group** |  |  |  |  |  |  |
| 11 years | 7.5% (4.7, 11.6) | 9.3% (5.7, 14.8) | 9.2% (5.4, 15.2) | 8.6% (5.7, 14.8) | 2.0% (0.6, 6.8) | 6.2% (3.2, 11.6) |
| 12 years | 2.7% (1.9, 4.0) | 4.0% (2.3, 7.0) | 3.4% (2.3, 5.0) | 2.5% (1.4, 4.4) | 0.7% (0.3, 1.6) | 1.2% (0.7, 1.8) |
| 13 years | 3.5% (2.9, 4.1) | 4.2% (3.0, 6.0) | 3.0% (2.4, 3.7) | 3.6% (2.7, 4.8) | 0.6% (0.4, 0.8) | 1.0% (0.8, 1.3) |
| 14 years | 4.9% (4.2, 5.8) | 5.2% (4.3, 6.2) | 5.1% (3.9, 6.5) | 4.7% (3.8, 5.8) | 1.1% (0.8, 1.6) | 2.0% (1.6, 2.4) |
| 15 years | 6.6% (5.5, 7.8) | 5.7% (4.9, 6.7) | 5.2% (4.4, 6.0) | 6.0% (4.4, 8.2) | 2.0% (1.2, 3.3) | 2.6% (2.1, 3.1) |
| 16 years | 8.4% (7.2, 9.7) | 8.0% (7.0, 9.0) | 8.2% (7.0, 9.7) | 5.9% (4.6, 7.6) | 2.7% (1.7, 4.1) | 3.8% (3.2, 4.5) |
| 17 years | 14.6% (13.0, 16.5) | 11.7% (10.3, 13.3) | 9.8% (8.7, 11.0) | 6.0% (4.4, 8.0) | 2.8% (1.7, 4.5) | 6.2% (5.4, 7.1) |
| **Gender** |  |  |  |  |  |  |
| Boys | 9.3% (8.1, 10.6) | 7.9% (6.7, 9.3) | 6.5% (6.0, 7.0) | 6.3% (5.3, 7.6) | 2.2% (1.5, 3.2) | 3.5% (3.1, 3.9) |
| Girls | 3.4% (3.0, 3.9) | 4.4% (3.8, 5.0) | 4.5% (3.8, 5.4) | 3.5% (2.7, 4.6) | 0.8% (0.5, 1.2) | 1.8% (1.5, 2.1) |
| **Overall prevalence** | **6.4% (5.8, 7.1)** | **6.2% (5.5, 7.0)** | **5.6% (5.1, 6.1)** | **5.2%(4.2, 4.9)** | **1.5% (1.1, 2.0)** | **1.9%(1.7 to 2.2)** |

*Notes:* 🞩 indicates that data is not available

**Supplementary Table 8: Robustness checks of the prevalence of use of any tobacco product, by country and African region(2010 – 2020)**

|  | **Use of any tobacco products** | | | |
| --- | --- | --- | --- | --- |
|  | **Boys (95% CI)** |  | **Girls (95% CI)** | **Total (95% CI)** |
| **African region** |  |  |  |  |
| Central Africa | 13.4% (10.7, 16.7) |  | 7.3% (6.0, 8.7) | 10.7% (8.7, 13.1) |
| East Africa | 10.6% (9.3, 12.1) |  | 6.4% (5.6, 7.4) | 8.8% (7.8, 9.8) |
| North Africa | 18.4% (15.3, 22.0) |  | 7.3% (4.9, 10.9) | 12.9% (10.6, 15.5) |
| Southern Africa | 32.0% (29.5, 34.6) |  | 25.0% (23.3, 26.7) | 28.8% (27.1, 30.6) |
| West Africa | 10.2% (8.9, 11.8) |  | 8.2% (6.3, 10.6) | 9.5% (8.0, 11.2) |
| **World Bank Income Group** |  |  |  |  |
| High income | 24.1% (20.9, 27.5) |  | 13.6% (11.2, 16.3) | 18.9% (16.5, 21.5) |
| Upper middle income | 36.3% (33.4, 39.4) |  | 26.6% (24.8, 28.6) | 31.5% (29.5, 33.6) |
| Lower middle income | 14.2 (12.5, 16.1) |  | 7.3% (5.9, 9.0) | 11.0% (9.8, 12.4) |
| Low income | 12.2% (11.0, 13.6) |  | 7.7% (6.6, 9.0) | 10.2% (9.3, 11.1) |
| **Age Group** |  |  |  |  |
| 11 years | 30.9% (23.2, 39.9) |  | 10.8% (6.0, 18.5) | 19.1% (14.4, 24.9) |
| 12 years | 11.6% (7.8, 16.9) |  | 5.8% (3.5, 9.6) | 8.2% (5.7, 11.6) |
| 13 years | 11.5% (9.2, 14.4) |  | 6.3% (5.0, 8.0) | 9.1% (7.7, 10.7) |
| 14 years | 13.6% (11.9, 15.5) |  | 10.6% (8.3, 13.3) | 12.2% (10.8, 13.9) |
| 15 years | 15.3% (13.4, 17.4) |  | 10.4% (8.8, 12.2) | 13.1% (11.6, 14.7) |
| 16 years | 21.7% (19.0, 24.6) |  | 13.1% (11.5, 14.9) | 17.9% (16.1, 19.7) |
| 17 years | 27.9% (25.1, 30.8) |  | 17.2% (14.8, 19.8) | 23.6% (21.6, 25.8) |
| **Overall prevalence** | 16.8% (15.5, 18.2) |  | 10.2% (9.0, 11.5) | 13.7% (12.7, 14.8) |

**Supplementary Table 9: Logit model marginal effects of the factors associated with the use of different products**

|  | (1) | (2) | (3) | (4) | (5) | (6) |
| --- | --- | --- | --- | --- | --- | --- |
| VARIABLES | Cigarettes | Other smoked tobacco | Smokeless tobacco | Shisha | Dual smokeless & smoked | Dual shisha & cigarettes |
| Age | -0.90444 | -1.1966*** | -0.4173 | 0.8238 | -3.5790** | 0.7814 |
|  | (0.61296) | (0.39970) | (0.5408) | (1.0318) | (1.4001) | (1.4149) |
| Age squared | 0.03891* | 0.0444*** | 0.01977 | -0.02046 | 0.1297** | -0.01099 |
|  | (0.02012) | (0.0139) | (0.01850) | (0.03494) | (0.0465) | (0.04846) |
| Female (Ref: Male) | -0.0386*** | -0.0280*** | -0.00485 | -0.0128** | -0.00793*** | -0.00595*** |
|  | (0.00385) | (0.00330) | (0.00329) | (0.00545) | (0.00260) | (0.00206) |
| *Region (Base: Central Africa)* |  |  |  |  |  |  |
| East Africa | -0.0292*** | -0.00558 | -0.0112** | -0.00282 | -0.00436 | -0.00201 |
|  | (0.0107) | (0.00416) | (0.00474) | (0.00530) | (0.00322) | (0.00315) |
| North Africa | -0.0193 | 0.00542 | -0.00440 | 0.0320*** | 0.000610 | 0.00178 |
|  | (0.0134) | (0.00632) | (0.00635) | (0.00842) | (0.00527) | (0.00368) |
| Southern Africa | -0.0253** | 0.00420 | -0.00286 | 🞩 | 0.00164 | 🞩 |
|  | (0.0118) | (0.00505) | (0.00535) | 🞩 | (0.00309) | 🞩 |
| West Africa | -0.0221* | 0.00928* | -0.0107** | 0.0167*** | 0.00159 | 0.00187 |
|  | (0.0126) | (0.00531) | (0.00509) | (0.00508) | (0.00489) | (0.00303) |
| *World Bank Income Group (Base: Low income)* | | | | | | |
| Lower middle income | -0.0212*** | -0.00207 | -0.00316 | 0.0112*** | 0.00423** | -0.000908 |
|  | (0.00559) | (0.00395) | (0.00466) | (0.00428) | (0.00196) | (0.00248) |
| Upper middle income | 0.0155* | 0.0215*** | -0.0110** | 0.00136 | -0.00153 | -0.00396 |
|  | (0.00883) | (0.00683) | (0.00539) | (0.0124) | (0.00294) | (0.00346) |
| Christian *(Base: Muslim & Hindu)* | 0.00189 | 0.00368 | -0.000655 | -0.00893*** | -0.00134 | -0.00182 |
|  | (0.00282) | (0.00230) | (0.00213) | (0.00259) | (0.00170) | (0.00123) |
| Age Restriction | 0.0476*** | 0.0308*** | 0.0280*** | 0.0554*** | 0.0172*** | 0.0184*** |
|  | (0.00401) | (0.00481) | (0.00417) | (0.00590) | (0.00275) | (0.00221) |
| Anti-tobacco use messages | -0.0119*** | 0.00597* | -0.000157 | 0.00192 | 0.00232 | -0.00171 |
|  | (0.00275) | (0.00353) | (0.00334) | (0.00603) | (0.00210) | (0.00187) |
| School teaching on tobacco use | -0.00481 | 0.00148 | 0.00497 | -0.0165*** | -0.000658 | -0.00430** |
|  | (0.00430) | (0.00329) | (0.00312) | (0.00457) | (0.00185) | (0.00181) |
| Health warnings | 0.0210*** | 0.0145*** | 0.0180*** | 0.0183** | 0.00423** | 0.00448** |
|  | (0.00464) | (0.00341) | (0.00343) | (0.00850) | (0.00215) | (0.00189) |
| Media exposure to smoking | 0.00192 | -0.00558 | -0.00547 | 0.00704 | -0.00431* | 0.00587*** |
|  | (0.00250) | (0.00350) | (0.00345) | (0.00636) | (0.00240) | (0.00196) |
| Tobacco adverts | -0.00317 | 0.00555 | 0.00269 | -0.00271 | 0.000682 | -0.00113 |
|  | (0.00458) | (0.00357) | (0.00345) | (0.00645) | (0.00200) | (0.00199) |
| Free tobacco product offer | 0.0184*** | 0.0468*** | 0.0487*** | 0.0359*** | 0.0191*** | 0.00756*** |
|  | (0.00538) | (0.00467) | (0.00387) | (0.00692) | (0.00396) | (0.00201) |
| Public exposure to smoking | 0.0243*** | -0.00237 | 0.00160 | 0.0117** | 0.00401** | 0.00865*** |
|  | (0.00425) | (0.00383) | (0.00320) | (0.00526) | (0.00182) | (0.00212) |
| School exposure to smoking | 0.0219*** | 0.0158*** | 0.0174*** | 0.00935** | 0.0121*** | 0.00682*** |
|  | (0.00456) | (0.00376) | (0.00342) | (0.00448) | (0.00228) | (0.00198) |
| Home exposure to smoking | 0.0281*** | 0.0219*** | 0.00681** | 0.0233*** | 0.00630*** | 0.00921*** |
|  | (0.00467) | (0.00384) | (0.00309) | (0.00495) | (0.00223) | (0.00175) |
| Peer smoking | 0.0664*** | 0.0354*** | 0.0351*** | 0.0441*** | 0.0172*** | 0.0208*** |
|  | (0.00429) | (0.00363) | (0.00391) | (0.00641) | (0.00275) | (0.00220) |
| Observations | 82,424 | 81,272 | 83,649 | 37,527 | 83,261 | 35,875 |

*Notes:* Robust standard errors are in parentheses. ***, **, and * indicates statistical significance at 1%, 5%, and 10%, respectively. The estimates associated with age and age squared are the raw coefficients from the Logit Model, not the Marginal Effects. We do not report the Marginal Effects for this variable since they are meaningless. Rather, the coefficients of age and age squared are taken together to determine the turning of age regarding the use of different tobacco products. 🞩 indicates that data was not available.
